# Clinically Translatable Uncertainty-Aware Machine Learning for Cardiovascular Disease Risk Analysis

**DOI:** 10.64898/2026.09.18.26363375

**Authors:** Sudharonjon Roy, Radiya Imran, Md Rejuan Haque, Fahad Mostafa

**Affiliations:** University of Alabama at Birmingham; Arizona State University; Ohio State University

**Keywords:** Cardiovascular disease, Machine learning, Causal association study, Uncertainty estimation, Explainable artificial intelligence, Precision medicine

## Abstract

**Importance:** Cardiovascular disease remains the leading cause of mortality worldwide. Although machine learning models have demonstrated strong predictive performance, limited attention has been given to uncertainty quantification and gender-specific risk factor identification, which are essential for clinically trustworthy artificial intelligence.

**Objective:** To develop and evaluate an interpretable and uncertainty-aware machine learning framework for cardiovascular disease prediction that integrates an association study on biomarkers, explainable artificial intelligence, and gender-stratified risk assessment.

**Design, Setting, and Participants:** This retrospective observational study utilized the UCI Heart Disease dataset comprising 918 participants, including 725 males and 193 females. Demographic, clinical, laboratory, and exercise-related variables were analyzed. Missing values were addressed using a hybrid imputation strategy combining MICE and supervised Random Forest imputation.

**Methods:** An ensemble machine learning imputation strategy was developed to address missingness. Then advanced machine learning models were developed and evaluated using stratified training/testing partitions and 5-fold cross-validation. Predictive performance was assessed using several evaluation metrics. Uncertainty was quantified through bootstrap resampling, predictive entropy, and mutual information. Linear Double machine learning and Causal Forest double machine learning were applied to estimate gender-specific averages and association studies for risk factors.

**Results:** Random Forest achieved the strongest and most stable predictive performance across male and female cohorts, with ROC-AUC values exceeding 0.90 and robust cross-validation performance. Hybrid imputation improved classification performance, increasing the F1-score from 0.87 to 0.89 and ROC-AUC from 0.90 to 0.92. The causal association study analyzes identified the number of major vessels, ST-segment slope, thalassemia, and exercise-induced angina as the strongest determinants of heart disease, with notable differences observed between male and female subgroups.

**Conclusions and Relevance:** Integrating predictive modeling, uncertainty quantification, and association study machine learning provides a robust and clinically interpretable framework for cardiovascular disease risk assessment. The proposed approach improves prediction reliability while identifying gender-specific causally important risk factors, supporting precision cardiovascular medicine and trustworthy clinical artificial intelligence.

## 1 Introduction

Recent estimates indicate that more than 500 million people are living with Cardiovascular Disease (CVD) globally, and in 2022 the disease accounted for approximately 20 million deaths, representing nearly one-third of all deaths worldwide. In the United States, CVD is the leading cause of death, responsible for about one in every three deaths annually [56]. Among cardiovascular conditions, coronary artery disease (CAD) represents a major cause of morbidity and mortality. Characterized by the narrowing or obstruction of the coronary arteries, CAD can reduce myocardial blood flow and lead to serious outcomes, including myocardial infarction, heart failure, and sudden cardiac death. Its burden is further amplified by the rising prevalence of lifestyle-related risk factors, including obesity, hypertension, diabetes, dyslipidemia, and smoking. Because CAD and other cardiovascular conditions develop silently over time, early detection and accurate risk prediction are crucial for timely intervention and effective prevention. Accordingly, identifying individuals at elevated risk of CVD has become a central objective in modern clinical research and preventive cardiology [13, 38, 45].

CVD risk is strongly influenced by age and sex and increases with age in both sexes due to the gradual deterioration of cardiovascular function and accumulation of risk factors. Males generally face higher risk and earlier disease onset compared to females, with coronary heart disease developing approximately 7–10 years earlier. In 2019, global age-standardized CVD mortality was higher in men than women, 280.8 vs. 204.0 deaths per 100,000, reflecting persistent sex disparities in cardiovascular outcomes. This disparity is associated with estrogen-related protection in premenopausal women, as well as sex-related differences in vascular structure and plaque formation [8].

CVD risk factors are commonly classified as non-modifiable and modifiable. Non-modifiable determinants—including age, sex, and genetic predisposition—establish baseline risk but cannot be changed. Modifiable factors—hypertension, smoking, poor diet, inactivity—are lifestyle-related and preventable. Research indicates that addressing these modifiable risk factors could substantially lower the incidence of CVD. However, early recognition of cardiovascular symptoms remains challenging, as symptoms such as chest discomfort, arm pain, weakness, dizziness, fatigue, and excessive sweating are often overlooked or attributed to other causes, resulting in delayed diagnosis and treatment. [5].

CVD risk is typically assessed using clinical tests and wearable sensing technologies that continuously monitor physiological signals, with results often recorded in electronic medical records (EMRs). However, EMR data are often unstructured, complicating risk factor identification, while wearable sensor data frequently contain noise and missing values, which compromises predictive accuracy. At the same time, CVD also imposes a substantial economic burden on patients, through increased healthcare costs, lost productivity, and reduced quality of life reflected in disability-adjusted life years. In the United States alone, the annual cost of CVD exceeds 200 billion, including both direct healthcare expenditures and indirect productivity losses [10]. These challenges highlight the need for intelligent systems capable of integrating and analyzing heterogeneous healthcare data to enable accurate and early prediction of CVD. With the growing availability of large-scale healthcare data, particularly electronic health records (EHRs), advanced analytical approaches have shown strong potential for identifying complex risk patterns and improving the prediction of CVD [41].

Traditional CVD risk prediction models for population-level risk stratification include the Framingham Risk Score, the Atherosclerotic Cardiovascular Disease (ASCVD) pooled cohort equations, and SCORE2 [16, 22]. However, these models are constructed using a limited set of demographic and laboratory variables, assume largely linear associations between predictors and outcomes, and typically rely on parametric regression frameworks. While these models do demonstrate acceptable discrimination within the different populations, their calibration can deteriorate across heterogeneous cohorts, such as underrepresented racial and ethnic groups, younger individuals, and patients with multimorbidity. Furthermore, these models do not account for complex physiologic interactions, stress-response dynamics, or subclinical markers of disease progression.

Recent advances in computational power and data availability have increased the application of machine learning (ML) methods to the prediction of cardiovascular risk. To address these challenges, ML methods, such as Random Forests (RF), Gradient Boosting machines (GBM), Support Vector Machines (SVM), and deep neural networks, are used in place of logistic regression to improve discrimination in select datasets [1, 15, 49]. These approaches can model nonlinear relationships, higher-order interactions, and complex dependencies among predictors without prespecified functional forms. These methods are useful because the growing availability of digitized EHRs, imaging repositories, wearable sensors, and genomic data has expanded the dimensionality for predictive modeling [14, 32].

Several studies have reported values of the area under the receiver operating characteristic curve (AUC) that exceed 0.85 for the detection of obstructive CAD or the prediction of major adverse cardiovascular events using ML techniques [29, 40]. Deep learning models applied to electrocardiograms have identified patterns predictive of reduced ejection fraction, atrial fibrillation, and incident mortality, even when such abnormalities are not apparent to expert clinicians [4, 42]. These findings suggest that ML may capture latent physiologic signals not readily apparent through traditional statistical modeling.

Despite these advances, transitioning ML models into routine cardiovascular practice remains a challenge. Many published studies rely on retrospective single-center datasets with modest sample sizes, raising concerns about overfitting and limited generalization. External validation is also frequently absent or incomplete, with calibrations—arguably as important as discrimination in clinical decision-making—inconsistently reported. In addition, algorithmic fairness and performance among demographic subgroups are rarely evaluated systematically. Lastly, complex “black-box” models often lack interpretability, creating barriers to clinician trust and regulatory approval [44, 53].

Interpretability is critical in cardiovascular medicine, where clinicians favor predictive tools grounded in established pathophysiology — such as ischemia-induced ST-segment depression, impaired chronotropic response, or angiographic burden. Explicit domain-informed feature engineering, including interaction terms between exercise-induced angina and ST depression or age-adjusted maximal heart rate, can enhance both model performance and transparency, bridging data-driven approaches with mechanistic plausibility. Sparse regularization techniques, such as the least absolute shrinkage and selection operator (LASSO), offer additional opportunities to enhance interpretability [55]. By penalizing model complexity and shrinking less informative coefficients toward zero, LASSO facilitates selection of a parsimonious subset of predictors. This approach not only mitigates overfitting but also supports derivation of simplified clinical risk scores. Historically, risk scores derived from Logistic Regression (LR) coefficients—scaled to integer points—have been instrumental in disseminating prediction tools at the bedside. Extending this paradigm within a modern ML framework may allow approximation of high-dimensional predictive performance while preserving usability.

Another emerging pillar of responsible medical AI involves explainability methods. Techniques such as SHapley Additive exPlanations (SHAP) decompose model predictions into feature-level contributions, enabling both global and local interpretability [9]. In cardiovascular applications, explainability reveals whether the models rely disproportionately on demographic variables, surrogate markers, or plausible physiological signals. The transparent explanation framework also facilitates auditing for bias and better aligns with evolving regulatory standards and reporting guidelines, such as CONSORT-AI and SPIRIT-AI [33, 43].

While ML models have shown strong performance in CVD classification, their predictions primarily reflect statistical associations rather than causal effects. Consequently, predictive importance does not necessarily indicate whether a clinical variable has an independent effect on disease risk. However, improvement in discrimination alone does not guarantee clinical utility. Even models with high AUC fail to meaningfully alter decision thresholds, risk reclassification, or downstream management. Decision-curve analysis, calibration assessment, and evaluation of net benefit are critical components of comprehensive model appraisal. Moreover, models must be assessed within the context of workflow integration, data availability, and potential unintended consequences, including exacerbation of health disparities.

In light of these considerations, there remains a need for evaluation of ML approaches to heart disease prediction that integrate feature engineering, interaction modeling, regularization-based selection, explainability, and translation into interpretable clinical scoring systems. Rather than treating ML and traditional statistical methods as competing paradigms, a complementary framework can leverage strengths of both: nonlinear modeling capacity from ML, and transparency and parsimony from regression-based risk assessment approaches.

To address these limitations, this paper applies Double Machine Learning (DML) to estimate the causal importance of key predictors while adjusting for confounding variables through flexible ML and orthogonalization to improve clinical interpretability [11]. This study introduces a sequential workflow for CVD prediction and risk factor identification. First, the heart disease dataset is preprocessed through missing-value handling, categorical encoding via an ensemble imputation strategy, and standardization of continuous variables. Multiple ML models are then developed, trained, and evaluated using train/validation/test splits and 5-fold cross-validation. Following this, predictive performance is assessed using F1-score, MCC, accuracy, ROC-AUC, AUPRC, and Brier score. Subsequently, uncertainty quantification is performed using bootstrap resampling and ensemble-based predictive uncertainty measures. Finally, in the association study, LinearDML and CausalForestDML were applied to estimate heterogeneous effects of clinically relevant risk factors, enabling individualized and interpretable CVD risk assessment.

## 2 Literature review

Since CVD remains the leading cause of death world-wide, many researcher have dedicated years to developing tools for CVD prevention, diagnosis, and management. Ali et al. [1] presented a smart healthcare monitoring system for heart disease prediction based on ensemble deep learning and feature fusion techniques. Their framework integrates physiological data collected from wearable sensors with EMRs to generate a comprehensive healthcare dataset. To improve predictive performance, the system employs information gain–based feature selection to remove irrelevant attributes and a conditional probability approach to assign class-specific feature weights. The final prediction is performed using an ensemble deep learning model enhanced with a LogitBoost meta-learning algorithm. Experimental results demonstrated that the proposed system achieved an accuracy of approximately 98.5%, outperforming several conventional ML classifiers. These findings suggest that combining heterogeneous healthcare data sources with ensemble deep learning can improve heart disease prediction performance. Korial et al. [30] proposed an ensemble-based system for early CVD detection by combining chi-square feature selection with a majority-voting classifier. Using 303 patient records and 13 clinical features from the Cleveland heart disease dataset, the chi-square method identified the five most informative features, reducing the computational burden by more than 50%. The voting ensemble integrated LR, RF, Gaussian Naïve Bayes (GNB), and *k*-nearest neighbors (KNN) classifiers and achieved an accuracy of 92.11%, representing a 2.95% improvement over the best-performing individual classifier. These findings demonstrate that combining feature selection with ensemble learning can improve both the predictive accuracy and computational efficiency of CVD detection. Wang et al. [55] conducted a retrospective comparative study developing risk prediction models for premature CAD using data from 797 patients, partitioned into training and validation sets at a 7:3 ratio. Both LASSO and RF models consistently identified hyperuricemia, chronic renal disease, and carotid artery atherosclerosis as the most important predictors. The RF model outperformed LASSO across all evaluation metrics, achieving an AUC of 0.91 compared to 0.84, with a statistically significant difference (Z = 3.99, P < 0.05). Based on SHAP-derived importance scores, carotid artery atherosclerosis, age, chronic renal disease, hyperuricemia, and gender were the highest contributing variables, leading the authors to conclude that RF offers greater clinical applicability for early screening and risk assessment of premature CAD. Dorraki et al. [15] examined the role of psychological factors in improving CVD prediction using an ensemble ML framework. Their approach combined five learning algorithms—DT, RF, XGBoost, SVM, and deep neural networks—within a majority voting ensemble model trained on UK Biobank data. When trained using only traditional cardiovascular risk factors, the ensemble model achieved an accuracy of 71.31%. However, incorporating psychological and mental health variables substantially improved predictive performance, increasing the accuracy to 85.13%. These findings highlight the importance of integrating psychological health indicators with conventional risk factors to enhance ML–based CVD prediction models. Olawade et al. [38] proposed a ML framework for CAD prediction integrating the Bald Eagle Search Optimization algorithm for feature selection to address the challenges of high-dimensional data and feature redundancy in medical datasets. Multiple classifiers were evaluated, including KNN, LR, and SVM with linear, polynomial, and RBF kernels, with RF consistently achieving the highest performance, attaining accuracies of 90% and 92% across the two datasets evaluated, when compared with the 71-73% accuracy achieved by traditional clinical risk scores. Key predictors identified included heart rate, age, BMI, systolic blood pressure, typical chest pain, ST elevation, and regional wall motion abnormality. The authors concluded that BESO significantly enhances feature selection efficiency while preserving predictive accuracy, and that ensemble-based methods offer superior generalizability for non-invasive coronary artery disease diagnosis. Dhanka et al. [13] developed a hybrid XGBoost-based framework for heart disease prediction. Their approach integrates statistical outlier detection techniques, including Z-score and interquartile range (IQR), together with hyperparameter optimization using the Optuna framework. The model was evaluated using the Cleveland dataset and demonstrated strong predictive performance, achieving an accuracy of approximately 95.45%, sensitivity of 92.86%, precision of 100%, specificity of 100%, and an F1-score of 96.3%. These results suggest that optimized gradient boosting techniques can improve the predictive performance of heart disease detection systems. Ali et al. [2] compared several supervised machine-learning algorithms for heart disease prediction using a publicly available dataset obtained from Kaggle. Their study evaluated KNN, DT, RF, and MLP classifiers and assessed feature importance for the applicable models. Among the evaluated algorithms, RF demonstrated the best predictive performance, achieving 100% accuracy, sensitivity, and specificity. The authors concluded that supervised machine-learning methods, particularly RF, may support accurate early-stage heart disease prediction and clinical decision-making. In a related study, Rabbi et al. [41] proposed ensemble-based ML approaches for heart disease prediction using multiple clinical datasets, including the Cleveland Heart Disease, Framingham Heart Disease, and Indicators of Heart Disease (2020) datasets. Their study introduced two ensemble models: a voting ensemble machine learning algorithm (VEMLA) and a bagging ensemble machine learning algorithm (BEMLA). The authors applied several classifiers, including RF, KNN, GNB, DT, LR, and SVM, combined with preprocessing techniques and dimensionality reduction methods such as principal component analysis (PCA) and linear discriminant analysis (LDA). Experimental results showed that VEMLA achieved an accuracy of 92% on the Cleveland dataset, while BEMLA achieved 97% accuracy on both the Framingham and Indicators of Heart Disease datasets, outperforming several individual classifiers. However, the reliance on specific benchmark datasets may limit the generalizability of the proposed approach to broader clinical settings. Talukder et al. [49] explored a hybrid explainable artificial intelligence-based machine learning framework (HXAI-ML) for CVD. The framework integrates advanced data balancing techniques, including Random Oversampling (RO), Synthetic Minority Oversampling Technique (SMOTE), Tomek Link (TL), and Instance Hardness Threshold (IHT), with ensemble ML classifiers such as RF, DT, GBM, and Extra Trees. To improve model interpretability, explainable AI tools including SHAP, LIME, and Permutation Importance Analysis (PIA) were incorporated to provide transparent insights into feature contributions. The HXAI-ML model achieved an accuracy of 98.78% and precision of 98.82% in CVD detection. Beyond prediction and interpretability, causal association study can also assess risk-factor relationships. Chernozhukov et al. [11] developed the Double/Debiased ML framework to address the regularization bias that renders standard estimators unreliable for causal inference. By employing the Neyman orthogonality condition alongside a cross-fitting procedure, the framework incorporates flexible algorithms such as Lasso, RF, and Neural Networks to handle high-dimensional confounding while keeping the target association parameter cleanly identified, yielding an estimator that achieves root-n consistency and asymptotic normality even when covariates substantially exceed the sample size. These results demonstrate the potential of integrating ensemble learning with explainable AI techniques to enhance both prediction accuracy and interpretability in clinical decision-support systems while indicating the need for complementary causal association study ML approaches.

From the above literature review, it can be observed that most existing studies primarily focus on improving CVD prediction accuracy through hybrid ML frameworks. Several investigations additionally explore feature engineering and feature selection techniques in combination with conventional classification models. Although these approaches have demonstrated promising predictive performance, relatively limited attention has been devoted to gender-specific risk factor estimation, uncertainty quantification, causally important predictors, and individualized treatment-effect estimation in cardiovascular ML research. In particular, few studies systematically evaluate the reliability and stability of model predictions through bootstrap-based uncertainty analysis, predictive entropy, or calibration assessment. Moreover, despite the increasing use of ML models for clinical risk stratification, the distinction between predictive association and causal importance of predictors remains insufficiently explored in CVD datasets. Existing studies rarely investigate heterogeneous treatment effects across clinically relevant subgroups such as gender and age, limiting the interpretability and translational applicability of current models. Therefore, there remains a need for an integrated framework that combines preprocessing procedures, such as hybrid missing-value imputation and class balancing, to improve model robustness in predictive modeling, uncertainty quantification, and causal DML approaches to enable robust, interpretable, and clinically translatable CVD prediction.

## 3 Materials and Methods

### 3.1 Heart disease dataset

The study utilized a structured clinical dataset derived from the Cleveland and Hungarian heart disease cohorts, sourced from the Heart Disease dataset [25] comprising demographic, physiological, and exercise-related variables routinely used in CVD assessment. The dataset includes key predictors such as age, gender, chest pain type, blood pressure, cholesterol levels, electrocardiographic findings, and exercise-induced responses. Several variables represent clinically established risk factors. For example, chest pain type characterizes symptomatic presentation, while maximum heart rate and exercise-induced angina reflect functional cardiac response to stress. Laboratory measures, including serum cholesterol and fasting blood glucose, capture metabolic risk. Structural indicators, such as the number of major vessels observed via fluoroscopy and thalassemia status, provide insight into CAD severity. The outcome variable encodes the presence and severity of heart disease. For classification modeling, this outcome variable was dichotomized to indicate the presence or absence of clinically significant disease. A detailed description of all variables, including definitions, types, and ranges, is provided in Table 1.

**Table 1:**
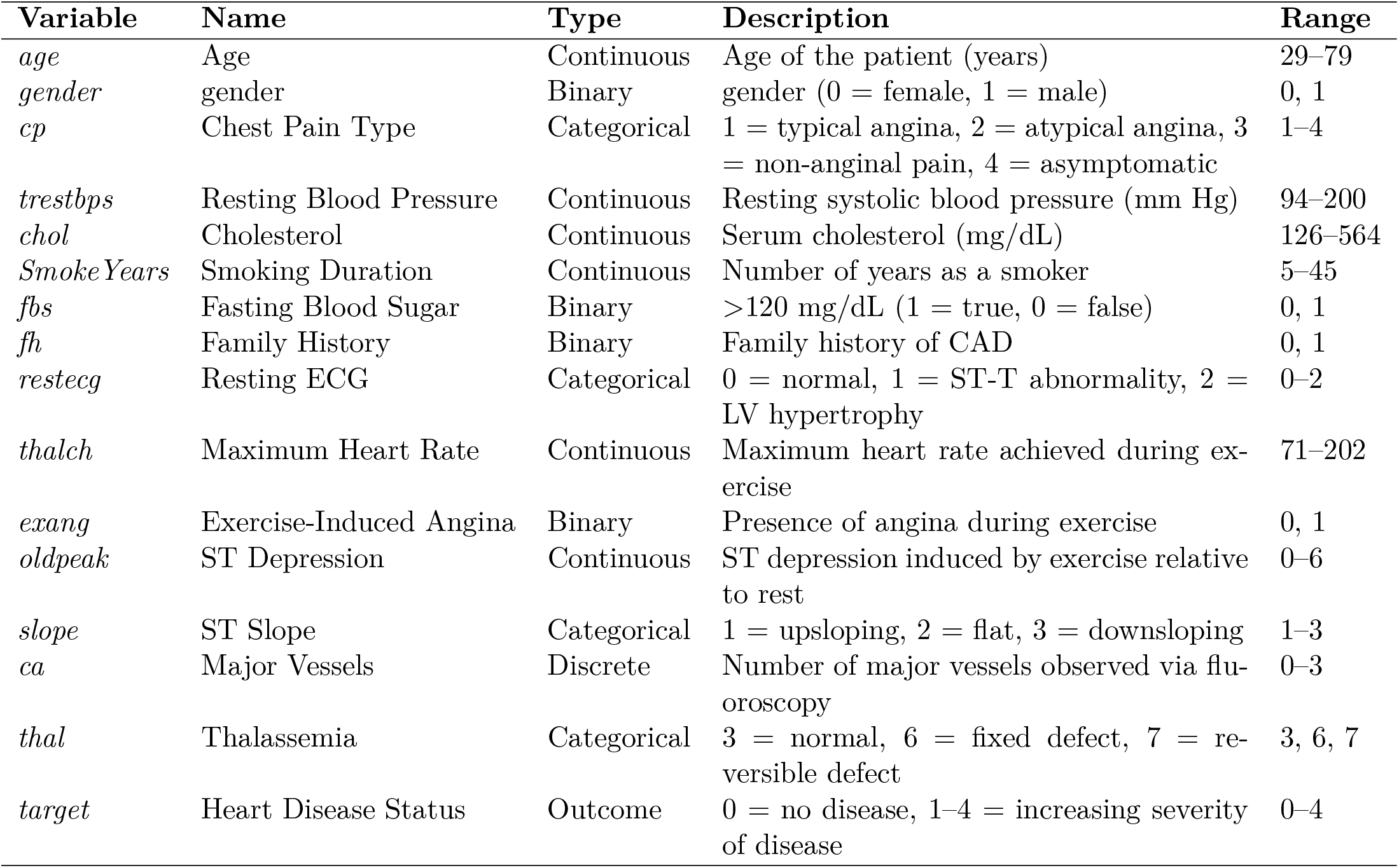
Clinical Variables Included in the CVD Prediction Model.

### 3.2 Study Design

This study employed a retrospective, observational design using publicly available clinical data to develop and evaluate ML models for CVD prediction, with particular emphasis on gender-stratified analysis and age-related differences. The methodological framework was structured around four integrated components: (i) standardized preprocessing within a unified column-wise pipeline to prevent data leakage; (ii) multi-algorithm variable selection using LASSO, Elastic Net, GBM, and RF to identify robust, model-agnostic predictors; (iii) predictive modeling using seven supervised classifiers spanning linear, margin-based, tree-based, probabilistic, instance-based, and neural paradigms—Logistic LR, SVM, DT, RF, GNB, KNN, and Multilayer Perceptron (MLP); and (iv) comparative performance evaluation across gender-stratified subgroups. The dataset was partitioned into training (80%) and testing (20%) subsets using stratified random sampling, and hyperparameters were tuned via stratified k-fold cross-validation on the training partition. Model discrimination and classification performance were assessed using the area under the receiver operating characteristic curve (auROC), area under the precision–recall curve (auPRC), F1-score, Matthews correlation coefficient (MCC), sensitivity, specificity, precision, and accuracy. This gender-stratified, multi-classifier framework enabled robust benchmarking of predictive performance, identification of subgroup-specific feature importance patterns, and assessment of model generalizability and reproducibility for clinically meaningful CVD risk stratification. The pipeline in Figure 1 includes exploratory data analysis, ensemble-based missing-value imputation and preprocessing, stratified training and testing with 5-fold cross-validation, development and evaluation of multiple ML classifiers, penalized and ensemble-based variable selection, gender-stratified risk factor analysis, bootstrap-based uncertainty quantification, and causal association analysis of CVD risk factors and their gender-specific effects.

**Figure 1:**
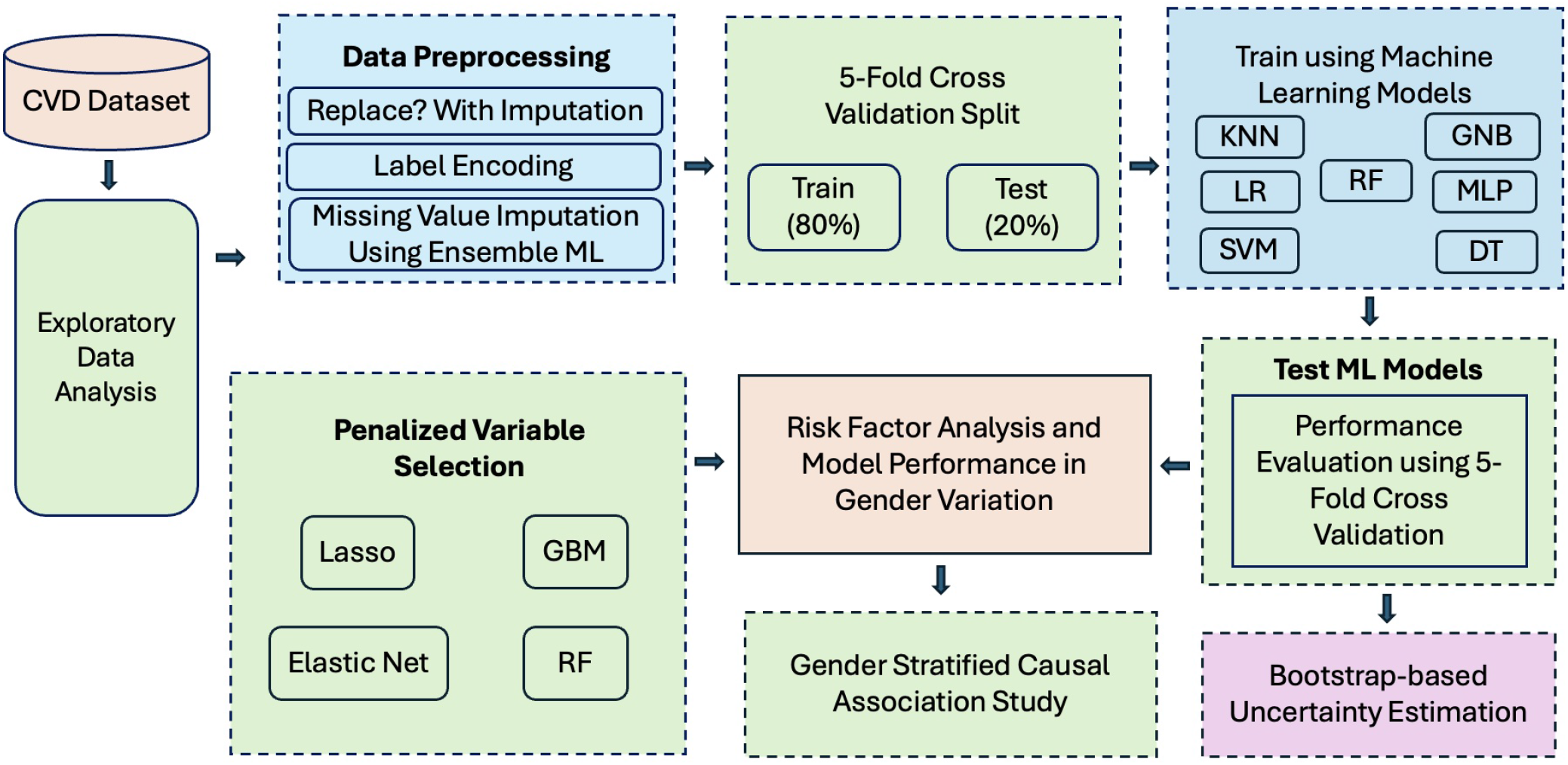
Study workflow for the proposed uncertainty-aware ML framework for CVD prediction with association study.

### 3.3 Data Preprocessing

The dataset consisted of structured clinical variables routinely used in CVD risk assessment, including demographic characteristics, laboratory measurements, and exercise-related indicators. The binary outcome variable indicated the presence (*Y* = 1) or absence (*Y* = 0) of heart disease. Prior to model development, a standardized preprocessing pipeline was implemented to ensure consistency across all ML models and to mitigate potential biases arising from heterogeneous feature scales and missing data.

### 3.4 Data Description

The dataset comprised clinical and demographic measurements collected for the purpose of heart disease classification. The full cohort included 918 participants, of whom 725 (79.0%) were male and 193 (21.0%) were female. The dataset represented a reasonably balanced proportion of patients with and without heart disease: 508 of the 918 participants (55.3%) were diagnosed with heart disease, while 410 (44.7%) were healthy controls. To investigate gender-specific patterns of heart disease, the data were stratified into male and female subgroups. Participants were further sorted into three age bands: 29–39 years (young adults; n = 80), 40–59 years (middle-aged adults; n = 735), and 60–77 years (older adults; n = 103). This stratification enabled the assessment of age and gender related differences in clinical risk factors and their associations with heart disease. Variables were grouped into four continuous predictors— serum cholesterol (*chol*, mg/dL), ST depression induced by exercise relative to rest (*oldpeak*), maximum heart rate achieved (*thalch*, bpm), and resting blood pressure (*trestbps*, mm Hg)—and eight categorical predictors: number of major vessels colored by fluoroscopy (*ca*), chest pain type (*cp*), exercise-induced angina (*exang*), fasting blood sugar *>* 120 mg/dL (*fbs*), resting electrocardiographic results (*restecg*), slope of the peak exercise ST segment (*slope*), thalassemia (*thal*), and the heart-disease outcome (*target*).

To examine the joint distribution of predictors across age and gender, continuous variables were displayed as kernel density estimates and categorical variables as frequency counts, stratified simultaneously by age category (columns) and gender (overlaid within each panel; Figure 2). The middle-aged subgroup accounted for the majority of observations across all categorical variables, consistent with the expected age distribution of heart disease cohorts, while the young-adult subgroup was sparsely populated and dominated by males. Continuous variables showed broadly similar central tendencies across age groups, with *thalch* shifting downward and *oldpeak* shifting upward in older adults, consistent with established age-related physiological changes. Categorical predictors revealed clear gender differences; most notably, males outnumbered females across nearly all levels of every categorical variable, and the disease outcome (*target*) was more prevalent in males within each age stratum. The marked imbalance between male (*n* = 725) and female (*n* = 193) participants motivated the gender-stratified modeling approach reported in subsequent sections.

**Figure 2:**
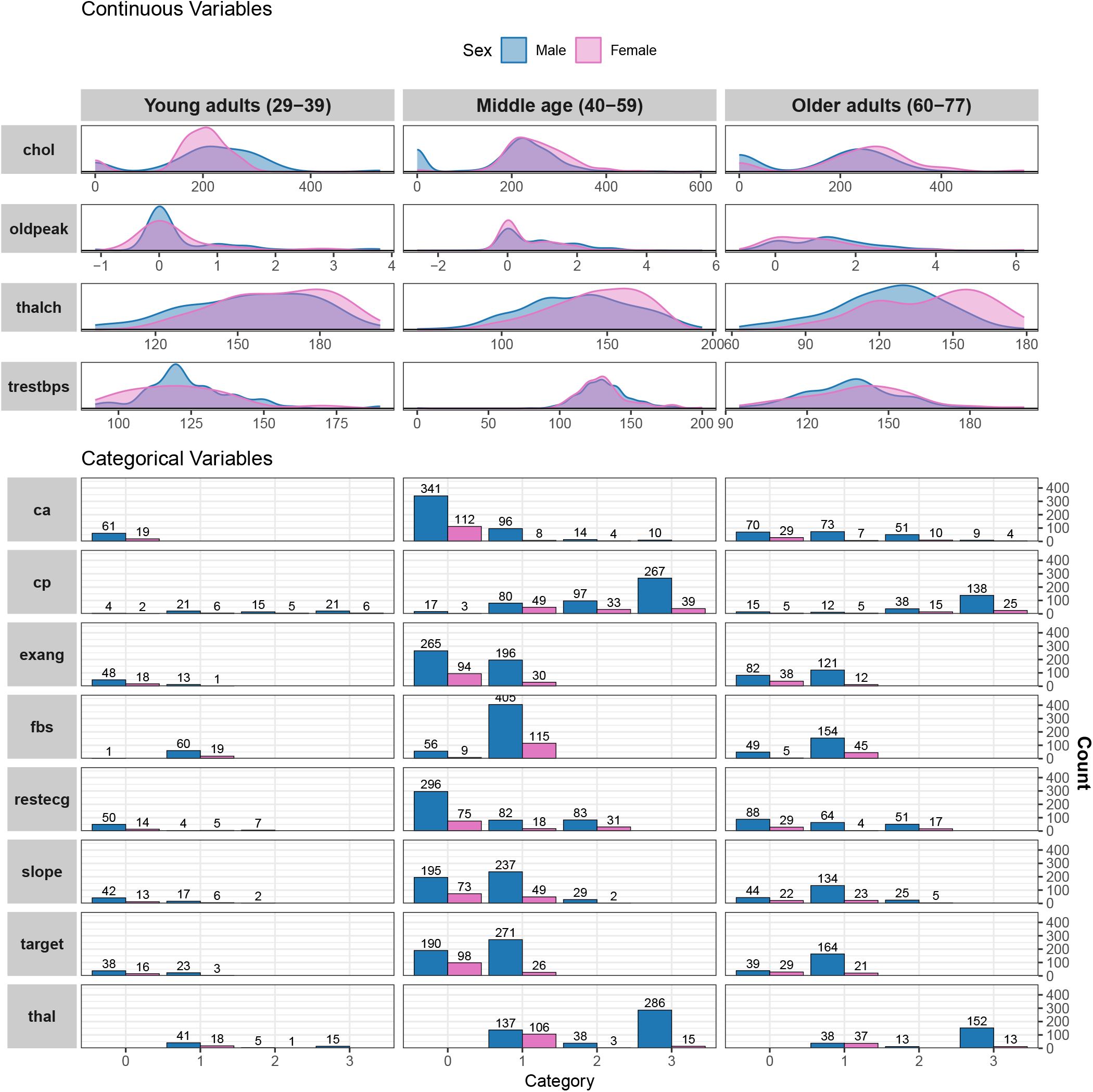
Distribution of continuous and categorical cardiovascular risk factors across age and sex groups. Continuous variables are shown as density plots to display their distributions, while categorical variables are shown as bar plots with frequency counts within each category.

### 3.5 Handling of Missing Data

Due to the complex nature of the CVD dataset, standard imputation techniques did not adequately address missing data, and can result in statistically insignificant outcomes. Therefore in the study, the missing data were addressed using a hybrid imputation strategy tailored to variable type. Continuous variables (e.g., age, resting blood pressure, cholesterol, maximum heart rate, and ST depression) were imputed using multiple imputation by chained equations (MICE) [36], allowing each variable to be modeled conditionally on all others. For categorical variables with missing values, a supervised imputation approach was applied. Specifically, for each categorical feature (e.g., gender, fasting blood sugar, exercise-induced angina) and multinomial features (e.g., chest pain type, electrocardiographic findings, number of vessels, and thalassemia), a RF regression was trained using observations with nonmissing values [50], incorporating all available predictors. Missing entries were then imputed using the predicted class labels. This approach preserves the discrete structure of categorical variables while leveraging multivariate relationships within the data, thereby improving the plausibility and consistency of imputed values. The full imputation algorithm is described in Algorithm 1.

### 3.6 Feature Scaling

All continuous predictors were standardized using z-score normalization:

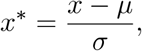

where *µ* and *σ* denote the sample mean and standard deviation of the data of each feature, respectively. Standardization was applied uniformly across all models to ensure comparability and numerical stability. While tree-based methods (e.g., RF and GBM) are scale-invariant, scaling is essential for regularized models such as LASSO and Elastic Net, where the magnitude of coefficients is directly influenced by feature scale [24].

### 3.7 Train–Test Split

The dataset was partitioned into training and testing subsets using an 80:20 split with a random seed of 42 in the Python scripts. Stratified sampling [48] was employed to preserve the proportion of positive and negative cases across subsets:

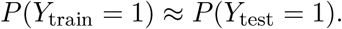

This is particularly important in clinical datasets where class imbalance may influence model performance. Moreover, to ensure methodological rigor and prevent data leakage [7], all preprocessing operations—including imputation, scaling, and encoding—were encapsulated within a unified column-wise transformation pipeline, wherein transformation parameters were estimated exclusively from the training set and subsequently applied to the test set. The preprocessing strategy was informed by both statistical and clinical considerations. Specifically, median imputation was employed to mitigate the influence of extreme physiological values, while standardization was applied to enhance numerical stability in optimization-based learning algorithms. Categorical variables of clinical significance, such as chest pain type and thalassemia, were transformed via one-hot encoding to preserve their nominal structure, given their established nonlinear associations with cardiovascular risk. Collectively, this integrated preprocessing framework promotes robustness, reproducibility, and interpretability in the development of predictive models for CVD.

### 3.8 ML Classification Model Development

Let 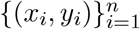 denote the heart disease male/female dataset, where *x*_*i*_ *∈* ℝ^*p*^ represents the vector of patient-level clinical features and *y*_*i*_ *∈ {*0, 1*}* indicates the absence or presence of CVD. The feature vector *x*_*i*_ includes demographic, physiological, and functional variables such as age, cholesterol levels, blood pressure, and exercise-induced angina, which are routinely used in clinical risk assessment. The objective is to learn a mapping function *ℱ* : ℝ^*p*^ *→* [0, 1] that estimates the conditional probability

#### Algorithm 1

Ensemble ML Imputation Strategy

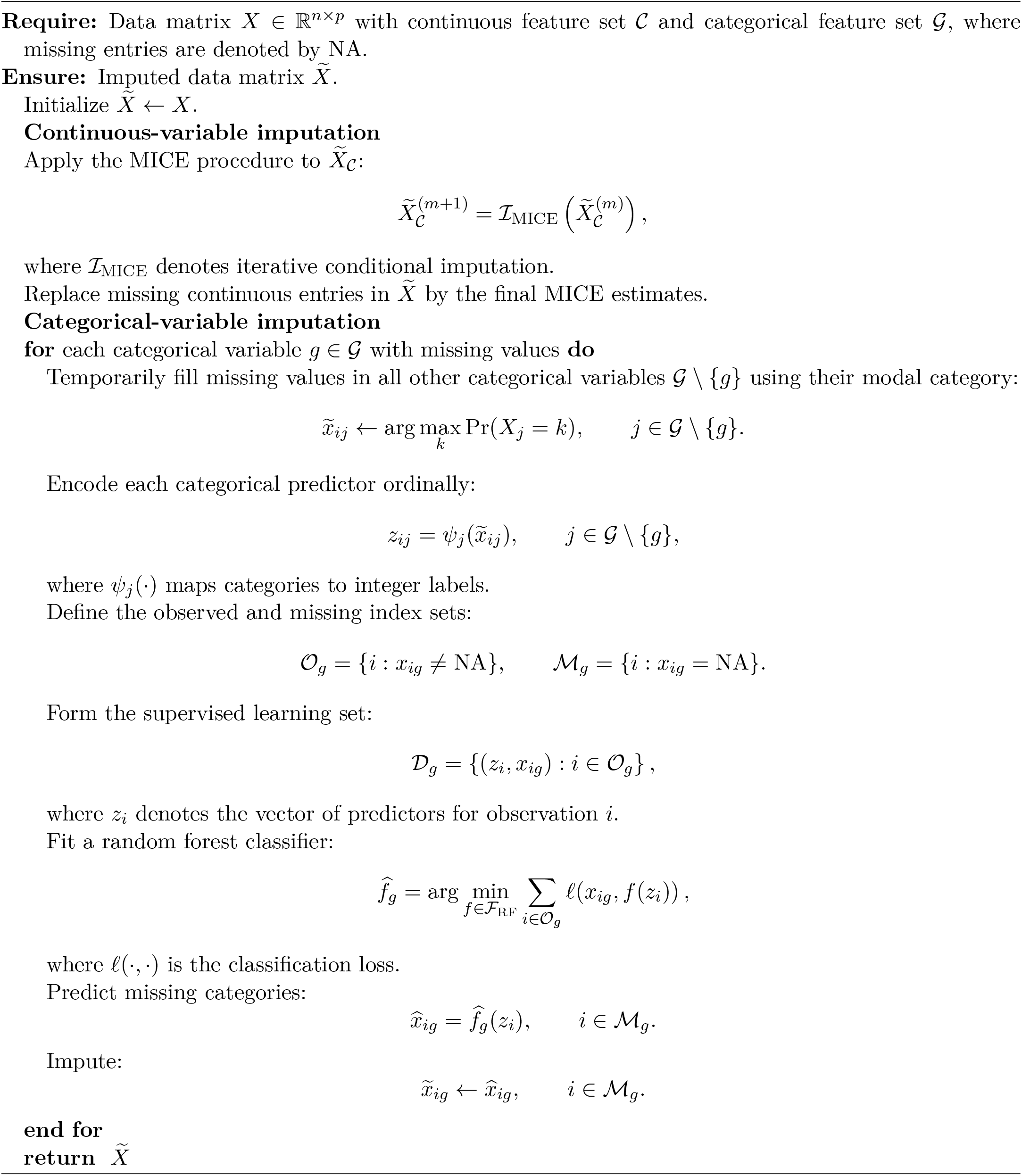

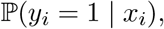

thereby enabling classification of patients into their predicted probability of low- and high-risk groups. Depending on the modeling framework, ℱ(·) may represent a linear predictor, a nonlinear transformation, or an ensemble of decision rules.

In this study, we consider a range of ML classification models, including both parametric and non-parametric approaches, to capture varying degrees of complexity in the relationship between clinical predictors and CVD outcomes. Linear models, such as LR, provide interpretable estimates of risk, while nonlinear methods, including tree-based ensembles and neural networks, allow for flexible modeling of interactions and complex dependencies among predictors. Each model is trained using the observed data 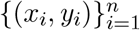 to approximate the underlying data-generating mechanism and to produce predictions for new patient profiles. The performance of these models is subsequently evaluated based on their ability to discriminate between diseased and non-diseased individuals, as well as their capacity to provide reliable probability estimates for clinical decision-making.

A diverse set of ML classification models was developed to evaluate CVD prediction using structured clinical variables. LR was implemented as a baseline probabilistic model because of its interpretability and ability to estimate clinically meaningful odds ratios [27, 41, 52]. SVM were used to model nonlinear decision boundaries through kernel-based transformations, enabling flexible separation of complex clinical patterns [3, 27, 41, 52]. DT were included because of their transparent hierarchical structure and clinically interpretable decision rules, while RF models were employed to improve predictive stability and reduce variance through ensemble learning [3, 27, 31, 41]. GNB classifiers provided a computationally efficient probabilistic baseline under conditional independence assumptions [3, 31, 41, 52]. In addition, the KNN algorithm was used to classify patients based on similarity in clinical feature space, thereby enabling patient-centric prediction [41, 52]. To capture higher-order nonlinear interactions among predictors, MLP neural networks were trained using layered nonlinear transformations [27, 31]. Finally, GBM were implemented to sequentially improve predictive performance by reducing errors from preceding weak learners [31, 41, 52]. Collectively, these models provide complementary perspectives on CVD prediction, ranging from interpretable linear frameworks to highly flexible nonlinear ensemble and neural network approaches.

### 3.9 Hyperparameter Optimization

Hyperparameter optimization was performed to improve model generalization and predictive stability across the different ML classifiers [17]. Model-specific tuning procedures were implemented using cross-validation on the training set to identify optimal parameter configurations while minimizing overfitting. For LR models, regularization parameters and penalty functions were optimized. In SVM, kernel parameters and margin regularization terms were tuned to improve nonlinear decision boundary estimation. Tree-based methods, including DT, RF, and GBM, were optimized with respect to tree depth, number of estimators, and node-splitting criteria to balance bias and variance. For KNN, the number of neighbors and distance metrics were evaluated, while MLP architectures were tuned by adjusting hidden layer structures, activation functions, and optimization parameters. Hyperparameter selection was guided primarily by cross-validated ROC-AUC performance, ensuring robust discrimination and stable model performance across CVD prediction tasks.

### 3.10 Penalized Variable Selection Strategy

To identify the most informative predictors of coronary heart disease we developed and evaluated four supervised ML architectures: LASSO [19, 21], Elastic Net [47, 57], GBM [13, 26], and RF [31, 38]. We selected LASSO and Elastic Net to manage potential multicollinearity and execute simultaneous regularization and variable selection. Concurrently, we leveraged the ensemble-based tree methods (GBM and RF) to capture complex, non-linear relationships and higher-order feature interactions without making prior structural assumptions about the data. To account for established gender-specific differences in cardiovascular pathophysiology, we stratified our entire cohort by biological gender and trained, tuned, and validated all predictive models independently within the male and female strata. This approach allowed derivation of distinct feature-importance profiles and coefficient pathways for each gender. We programmatically implemented the end-to-end analytical pipeline—including data preprocessing, hyper-parameter optimization via stratified k-fold cross-validation, and model evaluation—in Python (version 3.4) utilizing the scikit-learn (version 1.8) library.

#### 3.10.1 LASSO logistic regression

To identify the most relevant predictors of heart disease while avoiding overfitting, a LASSO was employed as the variable selection strategy. LASSO is well-suited for this purpose, as its *L*_1_ penalty automatically shrinks the coefficients of less informative predictors exactly to zero, effectively removing them from the model. The degree of penalization was governed by a single regularization parameter, which was optimized objectively through five-fold cross-validation on the training data, selecting the value that maximized the area under the ROC curve. The model was fitted using the SAGA optimization algorithm, with a maximum of 5,000 iterations to ensure convergence. All continuous predictors were standardized prior to fitting so that coefficients were on a comparable scale, and categorical variables were dummy-coded. Once the optimal regularization parameter was identified, the final model was refit on the complete training dataset using only the retained predictors. Variable importance was derived from the absolute magnitude of each predictor’s final coefficient, with multi-level categorical variables scored by summing the absolute coefficients across their indicator terms; all importance scores were then normalized to percentages summing to 100% to allow direct comparison across models [19, 21].

#### 3.10.2 Elastic Net logistic regression

Given the potential for multicollinearity among predictor variables, an Elastic Net was adopted to perform regularized variable selection for heart disease prediction. Unlike LASSO, Elastic Net combines both *L*_1_ and *L*_2_ penalties, improving model stability when predictors are correlated while retaining the ability to shrink irrelevant coefficients to zero. The mixing parameter was fixed at 0.5, assigning equal weight to both penalty components, and the overall regularization strength was optimized through five-fold cross-validation maximizing the area under the ROC curve. The model was fitted using the SAGA optimization algorithm with a maximum of 5,000 iterations. Variable importance was derived following the same procedure described for the LASSO model [47, 57].

#### 3.10.3 GBM Feature Importance

Beyond parametric penalization methods, a GBM was applied to assess variable importance through an ensemble-based approach. GBM is an ensemble method that sequentially combines shallow DTs, where each successive tree is fitted to the residual errors of the current ensemble, thereby minimizing the loss function through gradient descent in function space. The model was implemented using the GBM regressor in scikit-learn with a fixed random seed (number of random state = 42) to ensure reproducibility, using 100 boosting stages, a learning rate of 0.1, a maximum tree depth of 3, and a subsampling fraction of 1.0. The same preprocessing pipeline described for the LASSO model was applied. Variable importance was quantified as the cumulative reduction in node impurity attributable to each predictor across all trees, with multi-level categorical variables scored by aggregating importance values across their indicator terms, following the same normalization procedure described previously [13, 26].

#### 3.10.4 RF Feature Selection

Complementing the ensemble approach of GBM, an RF regressor was employed to further assess variable importance through an independent tree-based strategy. Unlike GBM, RF constructs multiple DTs in parallel, each grown on a bootstrap sample of the training data, with a random subset of predictors considered at every split to reduce correlation between trees and improve generalization. The model was implemented using the RF regressor in scikit-learn with 500 DTs (number of estimators = 500) and a fixed random seed (number of random state = 42) to ensure reproducibility, with all remaining hyperparameters retained at their scikit-learn defaults. The same preprocessing pipeline described for the LASSO model was applied. Variable importance was quantified as the mean decrease in Gini impurity attributable to each predictor across all trees, with multi-level categorical variables aggregated and normalized following the same procedure described previously [31, 38].

## 4 Evaluation Metrics

Model performance was evaluated using standard classification metrics [1]:

### Accuracy

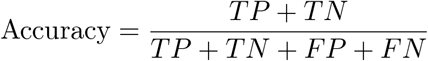

### Precision

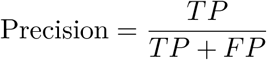

### Recall (Sensitivity)

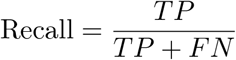

### F1-score

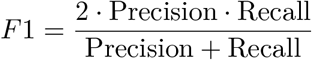

### Matthews Correlation Coefficient (MCC)

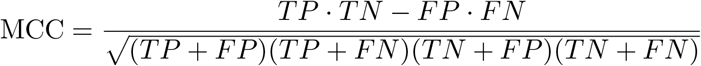

### ROC-AUC

The ROC-AUC measures the ability of the model to discriminate between classes across all decision thresholds:

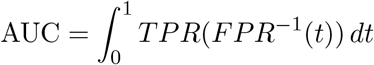

### Brier Score

The Brier score evaluates the accuracy of probabilistic predictions:

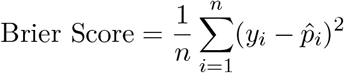

In the context of CVD prediction, sensitivity is critical for identifying high-risk patients, while ROC-AUC reflects overall discriminative performance. The F1-score balances precision and recall, particularly in imbalanced datasets. MCC provides a robust measure of classification quality even under class imbalance [51], and the Brier score assesses probabilistic calibration, indicating how well predicted probabilities correspond to observed outcomes.

## 5 Uncertainty Estimation for Performance Evaluation

Since in this study, *y*_*i*_ *∈ {*0, 1*}* indicates the absence or presence of CVD. The performance of the best-performing ML model was evaluated using accuracy, F1-score, MCC, ROC-AUC, AUPRC, and the Brier score.

To quantify uncertainty in performance estimates, bootstrap resampling was performed on the test set [23]. For each bootstrap replicate *m* = 1, … , *M* , performance metrics were recalculated:

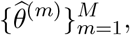

where 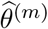 denotes the metric estimate from the *m*th bootstrap sample. Empirical 95% confidence intervals were obtained from the 2.5th and 97.5th percentiles of the bootstrap distribution.

Predictive uncertainty was additionally characterized using the variance of tree-wise probability estimates:

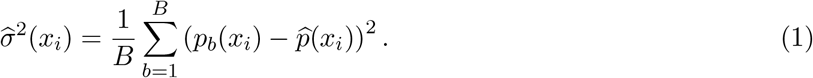

where *B* is the total number of trees, *p*_*b*_(*x*_*i*_) is the predicted probability for observation *x*_*i*_ from the *b*th tree, and

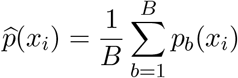

is the mean predicted probability across all trees.

Predictive entropy was computed as

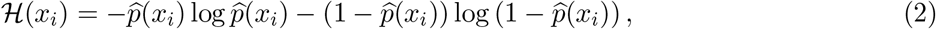

while epistemic uncertainty was quantified using mutual information:

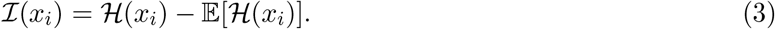

This framework enables simultaneous evaluation of predictive discrimination, calibration, and uncertainty in CVD risk prediction.

## 6 Methods on causal association study for analyzing biomarkers’ effects

### 6.1 LinearDML

The causal association effect of each clinical feature on heart disease probability was estimated using LinearDML [6, 11], a partially linear regression framework that isolates the average treatment effect (ATE) of each feature after flexibly adjusting for observed confounding via RF nuisance models. Each feature was treated as the treatment variable in turn, with the remaining features as covariates. Both the outcome and treatment were residualised on the covariate set through five-fold cross-fitting, and the ATE recovered by regressing outcome residuals on treatment residuals — yielding asymptotically valid confidence intervals and p-values. Models were estimated separately for male (n = 725) and female (n = 193) subgroups.

### 6.2 CausalForestDML

Heterogeneous treatment effects were estimated using CausalForestDML [6, 11, 54], a non-parametric extension of the DML framework that replaces the linear effect model with a causal forest, allowing treatment effects to vary flexibly across the covariate distribution. Each clinical feature was treated as the treatment variable in turn, with the remaining features as covariates. RF nuisance models and five-fold cross-fitting were used to residualise the outcome and treatment, after which the causal forest estimated individual-level conditional average treatment effects (CATEs). Reported ATEs represent the average of these CATEs within each gender-specific subgroup. This flexibility captures non-linear and subgroup-specific causal patterns that a linear model may miss, though at the cost of wider confidence intervals. Models were estimated separately for male (n = 725) and female (n = 193) subgroups. Under no unmeasured confounding, overlap, and partial linearity, ATEs represent the average causally important association contribution of each feature to heart disease risk [11]. ATEs were estimated using the LinearDML and CausalForestDML methods available in the EconML Python package [6].

## 7 Results

### 7.1 Missing Value Preprocessing

Figure 3 summarizes the missing data structure and the effect of mixed-type imputation using Algorithm 1 on model performance. Substantial missingness was observed in several clinically important variables, particularly *ca, thal*, and *slope*. These variables are strongly associated with CVD severity and therefore require careful handling to avoid biased estimation and loss of predictive information. Following the proposed hybrid imputation procedure, RF classification performance improved across all evaluation metrics. Specifically, the F1-score increased from 0.87 to 0.89, ROC-AUC improved from 0.90 to 0.92, MCC increased from 0.70 to 0.73, and AUPRC improved from 0.88 to 0.91. These findings suggest that supervised categorical imputation combined with MICE-based continuous imputation helped preserve important multivariate relationships within the data and reduced information loss caused by complete-case analysis. The improvement in MCC and AUPRC is particularly noteworthy, as these metrics are robust to class imbalance and provide a more comprehensive assessment of classification quality in clinical prediction settings. Overall, the results indicate that the proposed imputation framework enhanced both discrimination and calibration-related performance characteristics of the RF model.

**Figure 3:**
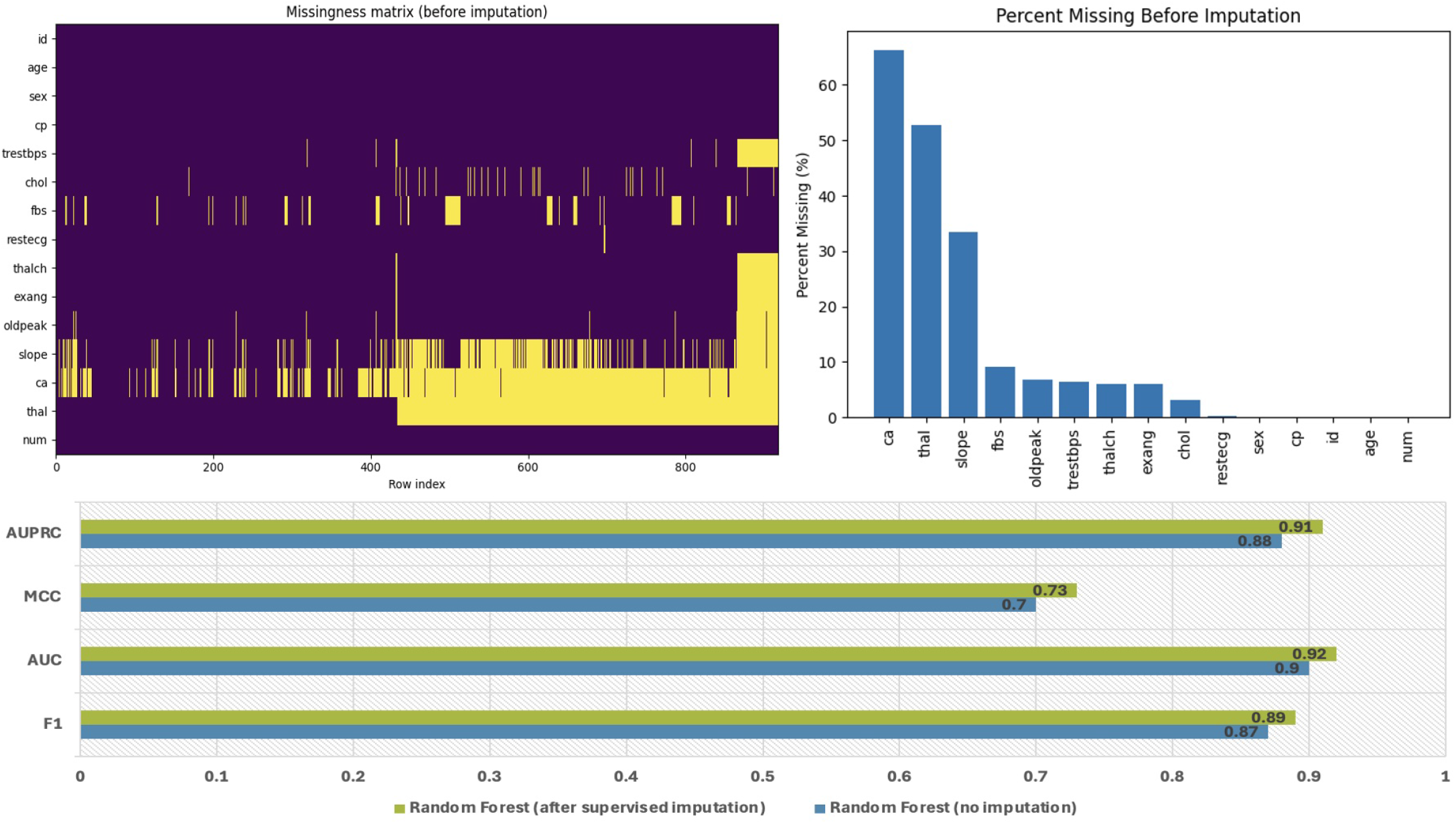
Missingness structure and comparison of predictive performance before and after supervised imputation in the heart disease dataset. The upper-left panel shows the missingness matrix, which shows substantial missingness in several variables. The upper-right panel presents the percentage of missing values for variables with the highest proportions of missingness. The lower panel compares RF classification performance before and after supervised imputation.

### 7.2 Descriptive Statistics

Age-stratified LR revealed a stronger association between age and heart disease among males than females (Table 2). Using adults aged 29–39 years as the reference group, males aged 40–59 years had significantly higher odds of heart disease (OR = 2.36, 95% CI: 1.37–4.14, *p* = 0.002), and the risk increased markedly among males aged 60–77 years (OR = 6.95, 95% CI: 3.76–13.1, *p <* 0.001). In contrast, age was not significantly associated with heart disease among females aged 40–59 years (OR = 1.41, 95% CI: 0.43– 6.41, *p* = 0.60), while females aged 60–77 years showed higher but borderline odds (OR = 3.86, 95% CI: 1.11–18.1, *p* = 0.051). These gender-specific differences may reflect biological and physiological variations in cardiovascular structure and regulation. Women generally have smaller hearts and coronary vessels and exhibit greater ventricular contractility, which leads to compensatory increases in heart rate. Additionally, estrogen provides cardioprotective effects in pre-menopausal women by improving vascular function and slowing the progression of atherosclerosis, thereby delaying the onset of CVD compared with males [8].

**Table 2:** Crude odds ratios (OR) and 95% confidence intervals (CI) for key predictors of heart disease, stratified by gender. Age group is also included with young adults (29-39 years) as the reference category.

| Characteristic | Male OR (95% CI) | Female OR (95% CI) |
| --- | --- | --- |
| N | 725 | 193 |
| cp | 3.01 (2.48, 3.69) | 4.44 (2.71, 7.83) |
| trestbps | 1.01 (1.00, 1.02) | 1.04 (1.02, 1.06) |
| chol | 1.00 (1.00, 1.00) | 1.00 (0.99, 1.00) |
| fbs | 0.51 (0.31, 0.81) | 0.23 (0.07, 0.70) |
| restecg | 1.23 (1.01, 1.49) | 1.16 (0.80, 1.68) |
| thalch | 0.96 (0.96, 0.97) | 0.98 (0.96, 0.99) |
| exang | 9.08 (6.26, 13.4) | 9.32 (4.42, 20.4) |
| oldpeak | 2.42 (2.00, 2.95) | 2.81 (1.90, 4.35) |
| slope | 6.80 (4.95, 9.45) | 8.71 (4.36, 19.2) |
| ca | 7.42 (4.94, 11.7) | 2.56 (1.64, 4.17) |
| thal | 3.07 (2.56, 3.71) | 6.36 (3.73, 12.3) |
| **Age group** |  |  |
| Young adults (29–39) | Reference | Reference |
| Middle age (40–59) | 2.29 (1.33, 4.04) | 1.41 (0.43, 6.41) |
| Older adults (60–77) | 6.76 (3.65, 12.8) | 3.86 (1.11, 18.1) |

### 7.3 Model Performance and Prediction Analysis

Figure 4 summarizes the predictive performance of multiple ML classification models for CVD prediction in male and female cohorts. Across both cohorts, ensemble-based approaches, particularly RF, consistently demonstrated strong predictive performance with high ROC-AUC, AUPRC, and F1-score values. LR also exhibited competitive performance, suggesting that clinically meaningful linear relationships remain highly informative for CVD risk stratification. In the female cohort, RF achieved among the highest overall discrimination performance, with test ROC-AUC and AUPRC values approaching 0.90 and strong cross-validation stability. LR and SVM also demonstrated robust performance, whereas DT and MLP models showed comparatively lower classification stability across several metrics. Similar trends were observed in the male cohort, where RF and LR achieved consistently high ROC-AUC and AUPRC values with relatively stable cross-validation estimates. Notably, GNB and KNN exhibited moderate predictive performance but greater variability across metrics. MCC values indicated improved classification balance for ensemble methods compared with simpler models, particularly in the male cohort. In addition, the similarity between test-set and cross-validation estimates across most models suggests limited overfitting and acceptable generalization performance. Ensemble methods, especially RF and GBM, demonstrated greater robustness in capturing nonlinear interactions among clinical predictors, while LR maintained strong interpretability and stable performance. Overall, these findings indicate that nonlinear ensemble approaches provide superior predictive discrimination for CVD classification, although simpler interpretable models such as LR remain competitive and clinically informative. The consistency of cross-validation results further supports the reliability and reproducibility of the proposed modeling framework across gender-stratified populations.

**Figure 4:**
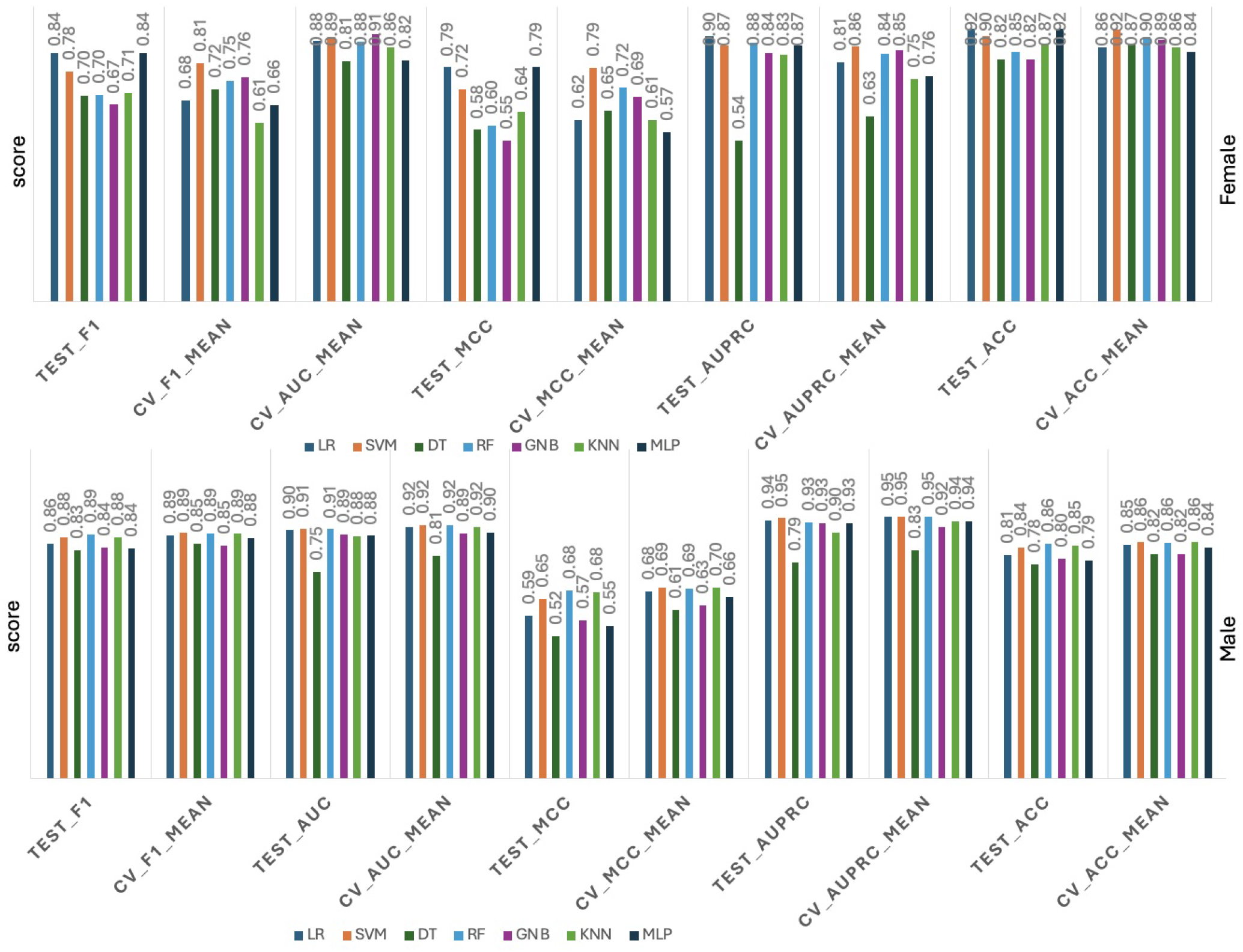
Comparative performance of ML classification models for CVD prediction in male and female cohorts.

### 7.4 Risk Factor Analysis and Model Performance in Gender Variation

#### 7.4.1 Male subgroup

Variable selection was performed using four classifiers—LASSO, Elastic Net, GBM, and RF—to rank predictors by their contribution to heart disease classification in the male subgroup (Figure 5). Across all four models, four variables consistently emerged as the most important predictors: *ca, cp, slope*, and *thal*. Notably, *ca* received the highest importance under both LASSO (*≈* 30%) and Elastic Net (*≈* 32%), while *cp* was the most important variable under RF (*≈* 17%) and *slope* dominated under GBM (*≈* 23%); *thal* ranked among the top four in every model (11–16%). Continuous predictors– age, *chol, oldpeak*, and *trestbps*—occupied intermediate ranks, while binary predictors (*restecg, fbs*) carried negligible weight in all four models. The convergence of categorical clinical and electrocardiographic predictors (*ca, cp, slope, thal*) at the top across multiple modeling frameworks indicates a robust, model-agnostic signal rather than an artifact of any single algorithm. These findings are consistent with the established cardiology literature: typical angina symptoms captured here by the *cp* variable are among the strongest independent predictors of obstructive CAD, with reported odds ratios of 2.5–3.5 in patients undergoing coronary angiography [37], and this association is particularly strong in men, who are more likely than women to present with classical anginal chest pain [20]. Likewise, men exhibit a substantially higher prevalence of obstructive multivessel and left main CAD than women, with reported odds ratios of approximately 2.7 for three-vessel disease and 7.4 for left main disease [28], which helps explain why *ca* carries such weight in the male model. Exercise-induced ST-segment changes—captured by *slope* and *oldpeak*—are well-established markers of inducible ischemia [18]. Together, these observations indicate that the predictors emerging from our male-specific models recapitulate the clinical and pathophysiological framework on which traditional risk stratification of CAD was largely built.

**Figure 5:**
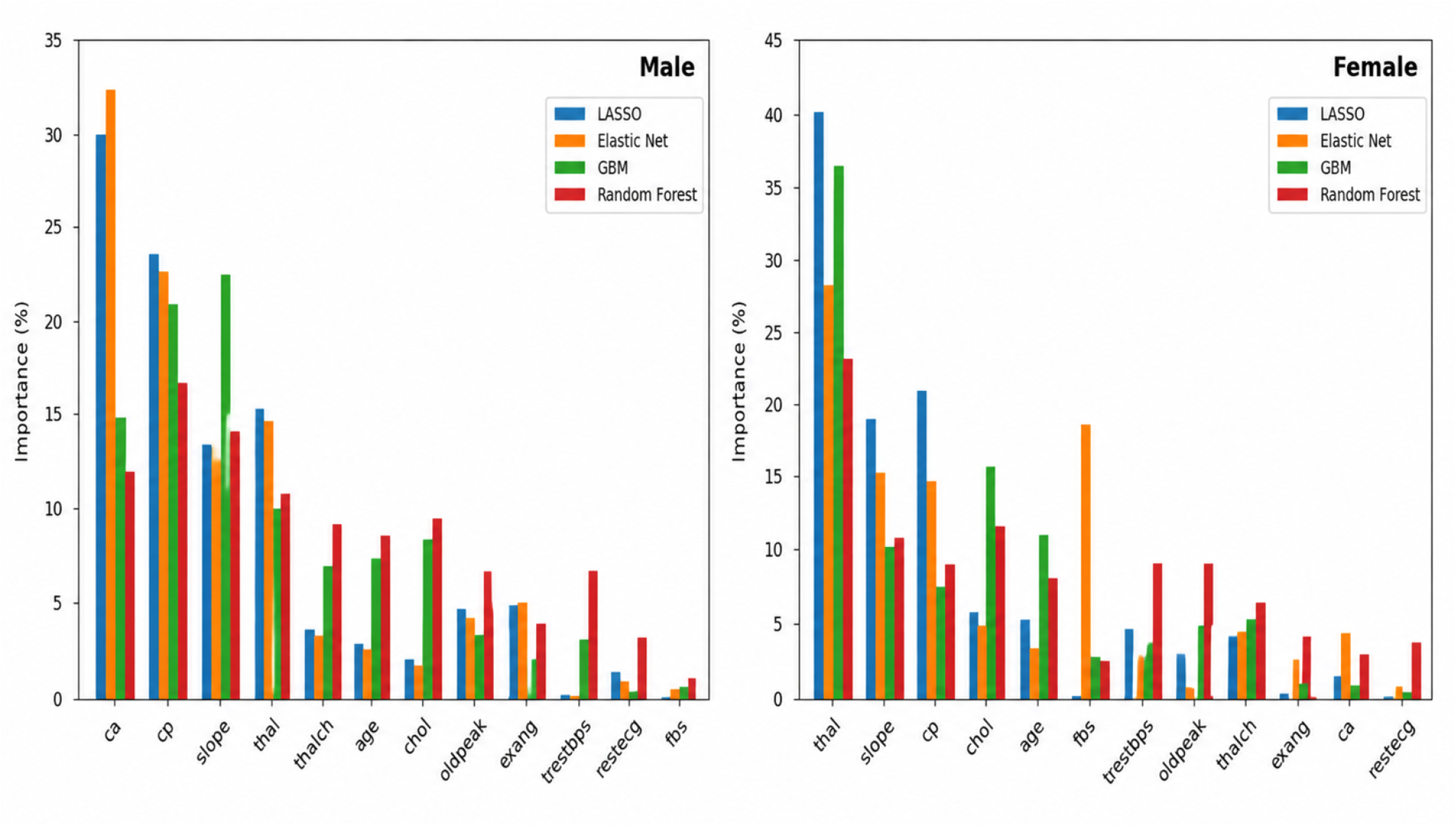
Feature importance ranking and comparison of CVD risk factors across four different ML models, indicated by different colors and stratified by gender.

Four ML–based feature selection approaches were evaluated using auROC, auPRC, sensitivity, specificity, and precision (Table 3). All approaches demonstrated comparable performance, with auROC values ranging from 90.4% to 91.6%. The RF–based feature selection approach achieved the highest au-ROC (91.6%), while the GBM approach produced the highest sensitivity (90.2%) and precision (87.4%). LASSO and Elastic Net feature selection methods yielded nearly identical performance (auROC 90.7% and 90.8%, respectively), indicating the stability and robustness of the selected features across different ML frameworks.

**Table 3:** Performance comparison of feature selection methods stratified by gender (%).

| gender | Model | auROC | auPRC | Sensitivity | Specificity | Precision |
| --- | --- | --- | --- | --- | --- | --- |
| Male | LASSO | 90.7 | 94.5 | 87.0 | 75.5 | 86.0 |
|  | Elastic Net | 90.8 | 94.5 | 84.8 | 75.5 | 85.7 |
|  | GBM | 90.4 | 94.2 | 90.2 | 77.4 | 87.4 |
|  | RF | 91.6 | 94.0 | 90.2 | 75.5 | 86.5 |
| Female | LASSO | 90.0 | 86.9 | 70.0 | 96.6 | 87.5 |
|  | Elastic Net | 93.8 | 90.5 | 80.0 | 96.6 | 88.9 |
|  | GBM | 93.1 | 84.4 | 80.0 | 89.7 | 72.7 |
|  | RF | 94.8 | 90.1 | 60.0 | 96.6 | 85.7 |

#### 7.4.2 Female subgroup

In the female subgroup, the variable importance rankings differed notably from those in males (Figure 5). Across all four classifiers, *thal* emerged as the strongest predictor, with importance ranging from approximately 23% under RF to 40% under LASSO—substantially higher than its weight in the male subgroup. *slope* and *cp* ranked second and third under most models (10–21%), maintaining an importance similar to that observed in males. The most striking gender-specific contrast was the elevated importance of serum cholesterol, which ranked among the top four predictors in GBM (*≈* 16%) and RF (*≈* 12%) for females but was only of modest importance in males. An additional notable finding was the prominence of *fbs* under Elastic Net (*≈* 19%), which was effectively zero in the male model and absent from the top predictors of the other three female models, suggesting that this association may be model-specific or driven by a small number of cases in the limited female sample. Conversely, *ca* —a top-three predictor in males across LASSO and Elastic Net—did not appear among the top ten in the female subgroup. These findings are consistent with the established literature on gender differences in CAD: Elevated cholesterol levels and LDL cholesterol increase after menopause in women, and the loss of estrogen’s protective cardiovascular effects may contribute to increased CVD risk compared with men [34]. Conversely, women are well documented to present more frequently with non-obstructive CAD on angiography than men, with approximately 60–70% of women undergoing coronary angiography for suspected ischemic heart disease showing non-obstructive disease compared with roughly 30% of men [12]. This pattern is reflected in lower vessel scores and a reduced burden of obstructive lesions among women [39], which provides a clinical explanation for why *ca* carried less predictive weight in the female subgroup. Together, these observations suggest that the relative contribution of clinical predictors to heart disease risk differs by gender, and that gender-stratified models may better capture clinically meaningful pathophysiological differences.

Model performance for the female subgroup using different ML–based feature selection approaches is summarized in Table 3. Overall performance was comparable to or slightly higher than that observed in the male subgroup, with auROC values ranging from 90.0% to 94.8%. The RF–based feature selection approach achieved the highest auROC (94.8%), followed by Elastic Net (93.8%) and GBM (93.1%). Sensitivity values ranged from 60.0% to 80.0%, whereas specificity remained consistently high (89.7%– 96.6%) across all approaches, indicating stronger performance in identifying healthy individuals than detecting positive cases. These findings may reflect the smaller female sample size and potential class imbalance, suggesting that interpretation of subgroup-specific performance should be made with caution.

### Comparison across gender

Comparing the two subgroups, several findings are worth highlighting. First, *cp, slope, thal, chol, oldpeak*, and *trestbps* appeared in the top ten for both genderes, indicating a shared core of predictors. Second, *ca* was uniquely important in males and *restecg* uniquely important in females, suggesting potential gender-specific biomarkers that merit further investigation. Third, although overall discrimination (auROC) was similar or higher in females, the trade-off between sensitivity and specificity differed substantially between gender—male models favored sensitivity, while female models favored specificity. Together, these results support the development of gender-stratified prediction models for heart disease, as a single pooled model may obscure clinically meaningful differences in predictor importance and operating characteristics.

### 7.5 Bootstrap-based Uncertainty Estimation on Performance

Figure 6 presents bootstrap-based uncertainty quantification (UQ) of RF model performance across male and female CVD cohorts. The distributions of F1-score, MCC, ACC, ROC-AUC, area under the precision–recall curve (AUPRC), and Brier score were estimated using repeated bootstrap resampling of the test set. Point estimates are shown alongside empirical 95% confidence intervals. Overall, the RF model demonstrated strong predictive performance in both cohorts, with relatively narrow uncertainty intervals for ROC-AUC and AUPRC, indicating stable discrimination performance. The male cohort exhibited consistently high predictive accuracy, with bootstrap distributions concentrated around high F1-score, MCC, and ROC-AUC values, suggesting robust classification stability across resampled datasets. The Brier score distribution was centered at comparatively lower values, indicating good probabilistic calibration. In the female cohort, the model also demonstrated strong discrimination, particularly for ROC-AUC and AUPRC, although broader bootstrap distributions were observed for F1-score and MCC compared with the male cohort. This increased variability likely reflects the smaller sample size and greater heterogeneity within the female subgroup. Despite this variability, confidence intervals remained within clinically acceptable ranges, suggesting preserved model reliability.

**Figure 6:**
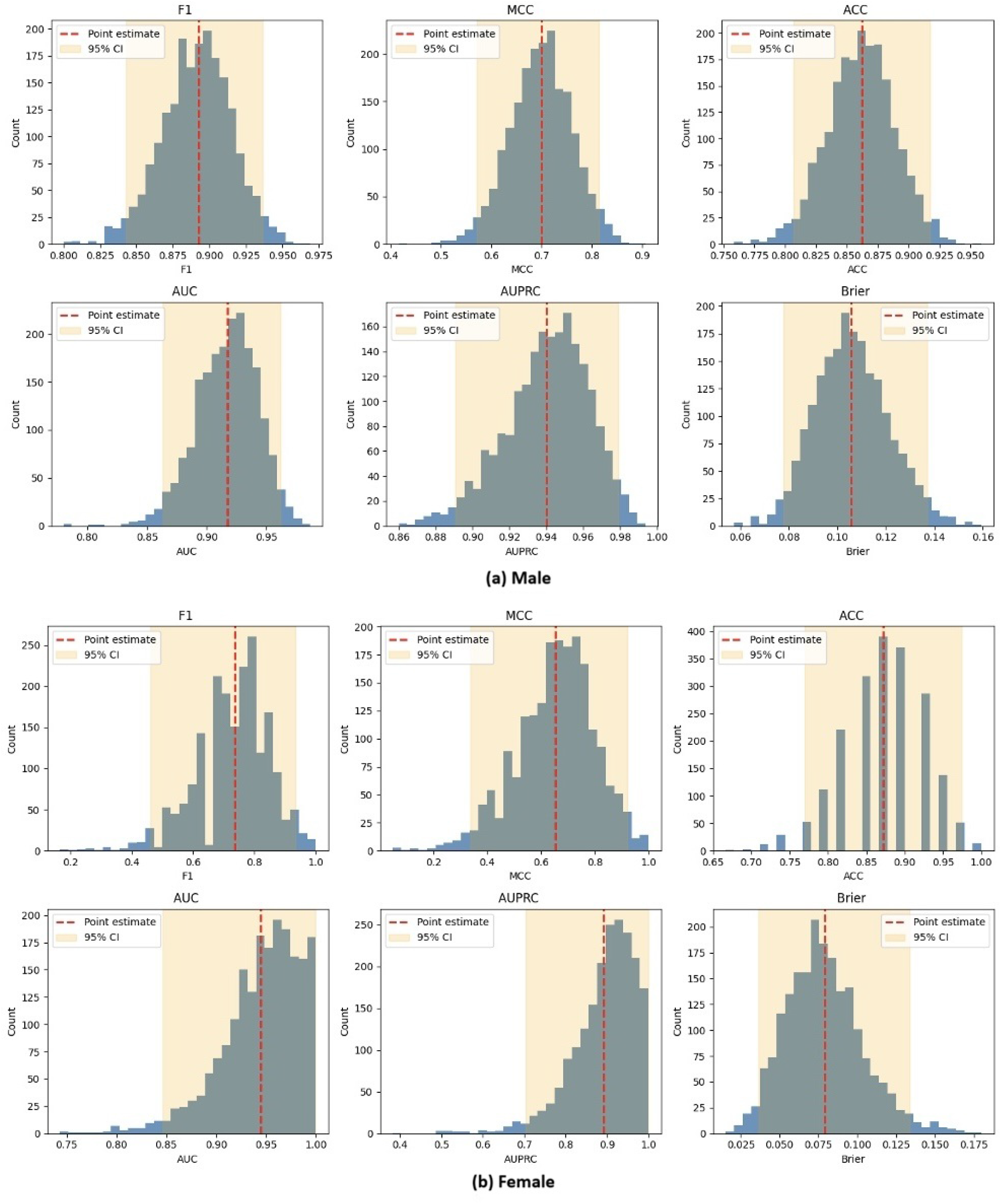
Bootstrap distributions and 95% confidence intervals of RF performance metrics stratified by gender.

Notably, ROC-AUC and AUPRC exhibited the smallest relative uncertainty across both cohorts, indicating that ranking-based discrimination was more stable than threshold-dependent classification metrics such as F1-score and MCC. The broader uncertainty observed in MCC and F1-score suggests sensitivity to threshold selection and class composition during resampling. In contrast, the relatively low and stable Brier scores support adequate calibration of predicted probabilities.

Collectively, these findings indicate that the proposed RF framework achieved strong and reproducible predictive performance while maintaining quantifiable uncertainty estimates across gender-stratified CVD populations.

### 7.6 Gender-specific Association study for analyzing biomarkers’ effects

Gender-stratified CausalForestDML and LinearDML analysis were applied to identify variables with robust, confounder-adjusted associations with heart disease across male and female subgroups. Rather than asserting causal directionality, this framework was employed to assess which predictors retain meaningful associations after accounting for the confounding structure of the observational data. As shown in figure 7, among males, *ca, slope, thal*, and *exang* demonstrated the strongest confounder-adjusted associations under both estimators, suggesting that structural coronary obstruction and ischemic abnormalities are robustly linked to heart disease risk beyond their correlational overlap with other features [12, 39]. Among females, *thal* and *slope* consistently retained significance across both estimators, confirming that ischemia-related pathways remain central to female cardiovascular risk; furthermore, *exang* was significant only under parametric LinearDML, possibly because the small female sample limited the precision of nonparametric CausalForestDML. Thus, its association was less robust than those of *thal* and *slope*. However, *ca* was non-significant in females, potentially reflecting gender differences in vascular pathology or reduced statistical power attributable to the lesser female sample (*n* = 193). Conventional risk factors — including age, cholesterol, resting blood pressure, and fasting blood sugar — showed negligible confounder-adjusted associations with confidence intervals spanning zero, indicating that their apparent predictive relevance in standard models may largely reflect shared correlational structure rather than independent associations. CausalForestDML produced wider confidence intervals than LinearDML throughout, consistent with its non-parametric flexibility, yet both estimators converged on the same significant features, strengthening confidence in these associations [6,54]. It is important to note that, as this analysis is based on observational data, unmeasured confounding cannot be ruled out, and these findings should be interpreted as confounder-adjusted associations rather than established causal effects [11]. Collectively, these results complement the variable importance findings by demonstrating that the top-ranked predictors are not merely spurious correlations, but rather robust associations that persist under rigorous confounding adjustment, underscoring the value of gender-stratified cardiovascular risk assessment.

**Figure 7:**
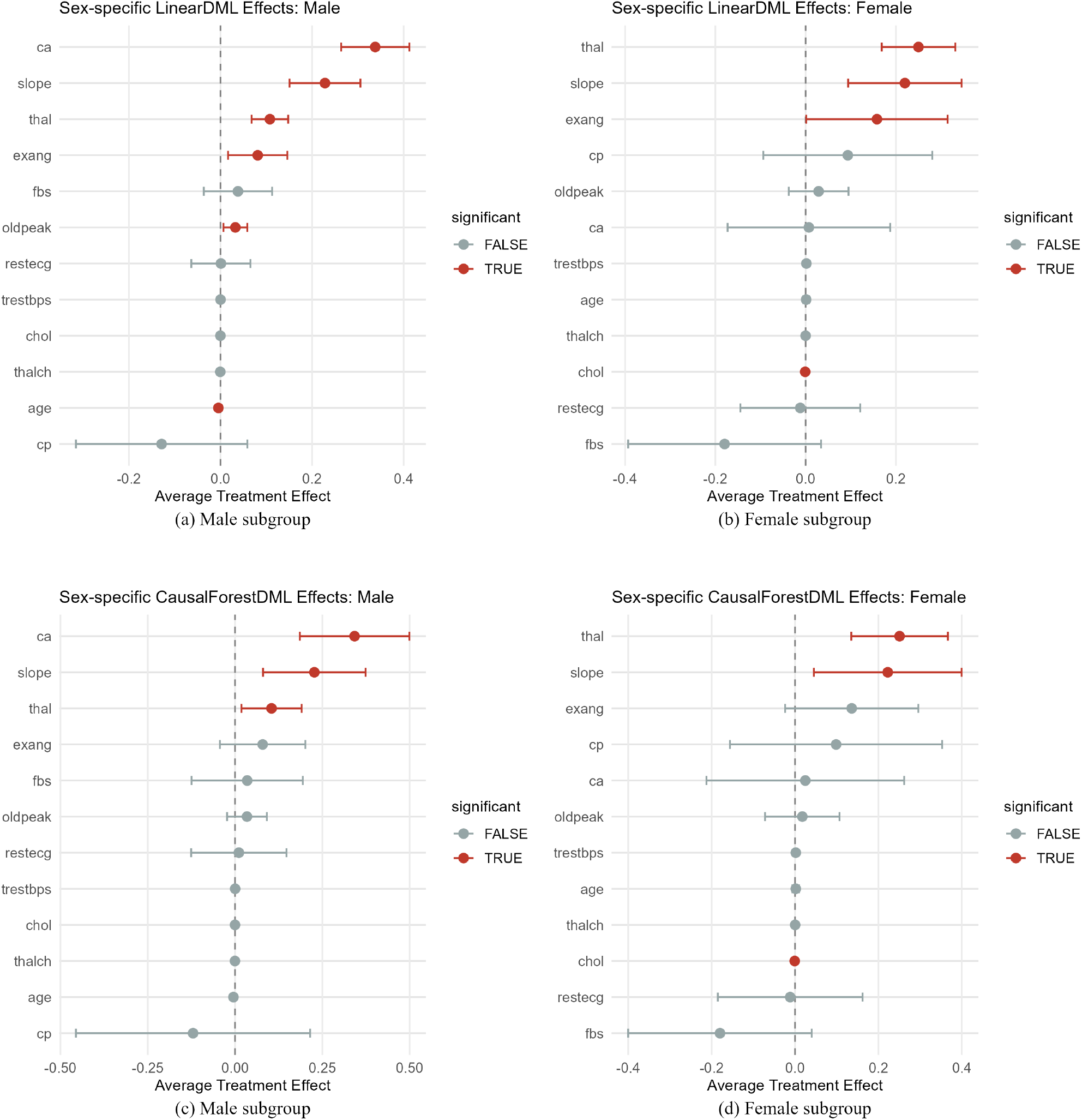
Gender-specific association study ML estimates of clinical feature effects on the probability of heart disease. Panels (a) and (b) show LinearDML estimates for male and female subgroups, respectively. Panels (c) and (d) show CausalForestDML estimates for male and female subgroups, respectively. Points represent average treatment effects, and horizontal bars represent 95% confidence intervals. Red points indicate statistically significant effects at the 5% level, while gray points indicate non-significant effects.

### 7.7 Calibration and Clinical Utility

Calibration curves demonstrated a generally good agreement between predicted probabilities and observed event rates across both male and female cohorts, indicating that the model reliably reflects true CVD risk. Accurate probability estimates are essential in clinical settings, as overestimation of risk may lead to unnecessary pharmacological intervention, while underestimation may result in missed opportunities for early preventive care [16, 22]. The bootstrap-estimated Brier scores illustrated in Figure 6, centered at approximately 0.10 in the male subgroup and 0.07 in the female subgroup, confirmed that the model produces reliable and well-calibrated probability estimates without systematically overestimating or underestimating CVD risk. The consistency between test-set and cross-validation performance estimates across most classifiers, particularly for ensemble methods such as RF, further reinforces the generalizability and reproducibility of the proposed modeling framework across gender-stratified populations. Furthermore, the gender-stratified modeling approach enhances clinical applicability by accounting for established differences in cardiovascular pathophysiology between males and females [8], ensuring that predicted risk estimates are tailored to each patient subgroup rather than derived from a single pooled model that may obscure clinically meaningful heterogeneity. Overall, these findings support the potential integration of the proposed framework into CVD screening and early detection workflows in clinical practice, although further external validation and clinical evaluation is needed before implementation [32].

## 8 Discussion

This study developed and evaluated an integrated artificial intelligence framework that combines ML, uncertainty quantification, explainable AI, and causal association parameter importance for CVD prediction using a clinically relevant heart disease cohort consisting of 918 participants, including 725 males and 193 females. The findings demonstrate that advanced ML approaches can achieve excellent predictive performance while simultaneously providing interpretable insights into disease mechanisms and individualized risk assessment. Importantly, the proposed framework addresses several limitations of previous studies [1,5,55] by incorporating uncertainty estimation, gender-stratified analyses, and association study ML, thereby improving the clinical trustworthiness and translational potential of AI-driven cardiovascular risk prediction.

A major finding of this study is the substantial improvement obtained through the proposed hybrid imputation strategy. Several clinically important variables exhibited considerable missingness, particularly *ca, thal*, and *slope*. Following imputation, RF performance improved across all evaluation metrics, with the F1-score increasing from 0.87 to 0.89, ROC-AUC improving from 0.90 to 0.92, MCC increasing from 0.70 to 0.73, and AUPRC increasing from 0.88 to 0.91. These results suggest that the combination of MICE-based continuous imputation and supervised RF categorical imputation effectively preserved important multivariate relationships within the data while reducing information loss associated with complete-case analysis [50].

Among the evaluated classifiers, RF emerged as the strongest overall performer. In the male subgroup, RF achieved the highest auROC (91.6%) while maintaining strong sensitivity (90.2%), specificity (75.5%), precision (86.5%), and auPRC (94.0%). In the female subgroup, RF again achieved the highest auROC (94.8%), although the sensitivity was lower (60.0%) and specificity remained high (96.6%). These findings indicate that ensemble methods are particularly effective in capturing the complex nonlinear relationships that characterize CVD [35]. Notably, LR remained highly competitive, achieving auROC values of 90.7% and 90.0% in males and females, respectively, suggesting that clinically meaningful linear associations continue to provide substantial predictive information. The close agreement between test-set and cross-validation performance further supports the robustness and generalizability of the proposed modeling framework.

The feature importance analyses revealed both shared and gender-specific cardiovascular risk factors. In males, *ca, cp, slope*, and *thal* consistently ranked among the most important predictors across all ML methods. The importance of *ca* reached approximately 30% under LASSO and 32% under Elastic Net, while *slope* contributed approximately 23% under Gradient Boosting and *cp* approximately 17% under RF. In contrast, the female subgroup demonstrated a different risk profile, with *thal* emerging as the dominant predictor, contributing as much as 40% under LASSO and approximately 23% under RF. Furthermore, cholesterol showed substantially greater importance in females than males, while *ca* lost much of its predictive contribution. These observations are consistent with known gender-specific differences in cardiovascular physiology and CAD presentation [8, 28].

The uncertainty quantification analyses further strengthen the reliability of the proposed framework. Bootstrap resampling demonstrated relatively narrow confidence intervals for ROC-AUC and AUPRC in both gender, indicating stable discrimination performance. The male cohort exhibited concentrated distributions of F1-score, MCC, and ROC-AUC, whereas broader uncertainty distributions were observed among females, likely reflecting the smaller female sample size and increased heterogeneity. Nevertheless, calibration remained strong in both groups, as indicated by consistently low Brier scores and stable uncertainty intervals. These findings emphasize the importance of reporting predictive uncertainty alongside conventional performance metrics when deploying ML models in clinical settings [23].

### Strengths, Limitations, and Validation

One of the most important contributions of this work is the integration of association study ML into the assessment of cardiovascular risk. Gender-stratified DML analyses identified *ca, slope, thal*, and *exang* as the strongest associations contributing to heart disease risk among males. In females, *thal* and *slope* remained significant, whereas *ca* became non-significant, suggesting important biological and pathophysiological differences between gender. Interestingly, several traditional cardiovascular risk factors, including age, cholesterol, resting blood pressure, and fasting blood sugar, demonstrated negligible average treatment effects after adjustment for confounding. This distinction between predictive association and causal contribution highlights the value of causal association study ML for identifying genuinely actionable risk factors [11].

Despite these strengths, several limitations should be acknowledged. The analyses were conducted using retrospective observational data from publicly available cohorts, and external validation using larger multi-center populations remains necessary. The smaller female subgroup should also be considered when interpreting the gender specific findings. Given the established differences in cardiovascular risk factors and disease presentation between males and females, future studies should validate these results using a larger, more representative population [8], and evaluate additional factors relevant to cardiovascular health. These include mental health conditions such as depression, anxiety, and psychosocial stress, which have been associated with adverse pregnancy outcomes and cardiovascular health in women [46]. Additionally, causally feature importance association investigation relies on assumptions regarding measured confounding and overlap that cannot be fully verified in observational datasets [11]. Future research should investigate external validation, prospective clinical deployment, integration of EHRs and wearable sensor data, and longitudinal modeling of cardiovascular risk trajectories [32]. Further studies should also assess the generalizability and practical applicability of the proposed framework across different populations and healthcare settings before broader adoption in clinical or public health settings.

### Public Health Implementation

From a public health perspective, these findings have several important implications. First, the ability to identify high-risk individuals with high predictive accuracy may facilitate earlier intervention and targeted preventive care, potentially reducing cardiovascular morbidity and mortality. Second, the identification of gender-specific causal determinants supports the development of personalized prevention strategies and precision cardiology initiatives. Third, uncertainty-aware predictions may improve clinician confidence and promote responsible integration of AI systems into routine healthcare workflows, as uncertainty estimates can assist clinicians in identifying predictions that require further evaluation or careful interpretation [9, 44, 53]. Finally, causal identification of modifiable risk factors such as exercise-induced ischemic responses may support more effective allocation of public health resources toward screening, prevention, and lifestyle interventions. ML can augment, though not replace, established cardiovascular risk prediction by integrating multiple risk factors and identifying patterns in the data, although wider clinical use will depend on prospective validation in real-world settings first [16, 32].

## 9 Conclusion

CVD remains a major global health challenge, highlighting the need for prediction models that are accurate, reliable, and clinically interpretable. This study developed an integrated artificial intelligence framework that combines hybrid data preprocessing, advanced ML, uncertainty quantification, explainable AI, and causally feature importance association study for CVD prediction using structured clinical data. A hybrid imputation strategy incorporating MICE for continuous variables and supervised RF imputation for categorical variables improved data quality and enhanced predictive performance, increasing the F1-score from 0.87 to 0.89 and ROC-AUC from 0.90 to 0.92. Among the evaluated ML models, RF consistently achieved the strongest overall performance across both male and female cohorts, producing ROC-AUC values exceeding 0.90 together with high AUPRC, F1-score, MCC, and good calibration while maintaining stable generalization under cross-validation. Bootstrap-based uncertainty quantification further demonstrated that model predictions were robust and reproducible, providing confidence in the reliability of the proposed framework for clinical risk prediction. A major contribution of this study is the integration of causal ML into cardiovascular risk assessment. LinearDML and CausalForestDML distinguished causal determinants from predictive associations, consistently identifying the number of major vessels, ST-segment slope, thalassemia status, and exercise-induced angina as the strongest contributors to heart disease risk. Furthermore, the observed gender-specific differences emphasize the importance of personalized cardiovascular risk assessment rather than relying on a single population-wide prediction model. Overall, the proposed framework demonstrates that combining high-performing ML with uncertainty quantification and causal predictor importance association study provides a robust, interpretable, and clinically translatable approach for CVD prediction. Future studies should validate the framework using large multi-center cohorts, longitudinal EHRs, and prospective clinical implementation to support precision cardiovascular medicine and trustworthy clinical artificial intelligence.

## Data Availability

A. Janosi, W. Steinbrunn, M. Pfisterer, and R. Detrano. Heart disease dataset. UCI Machine Learning Repository: https://archive.ics.uci.edu/dataset/45/heart+disease

## Acknowledgments

The authors are thankful to the ASU Supercomputing Center-SOL and the writing center for their support.

## Authors Contribution

Authors SR, and FM contributed to the formulation and analysis of the models, graphical presentations, data curation, and the simulation study; FM conceptualized and directed the research. SR, RI, MRH, and FM contributed to the literature search, model evaluations, and the writing and editing of the article.

## Code, Data and supplementary material availability

Will be provided on the request.

## Ethics Approval

There is no ethical approval needed due to the use of simulated and publicly available data.

## Clinical Trial Registration

The authors did not use clinical trial data directly. The authors used publicly available data with proper references in the text.

## Generative AI statement

The author(s) declare that no Generative AI was used in the creation of this manuscript.

## References

[1] F. Ali, S. El-Sappagh, S. R. Islam, D. Kwak, A. Ali, M. Imran, and K.-S. Kwak. A smart healthcare monitoring system for heart disease prediction based on ensemble deep learning and feature fusion. Information Fusion, 63:208–222, 2020.

[2] M. M. Ali, B. K. Paul, K. Ahmed, F. M. Bui, J. M. Quinn, and M. A. Moni. Heart disease prediction using supervised machine learning algorithms: Performance analysis and comparison. Computers in biology and medicine, 136:104672, 2021.

[3] M. U. Aslam, S. Xu, S. Hussain, M. Waqas, and N. L. Abiodun. Machine learning-based classification of valvular heart disease using cardiovascular risk factors. Scientific Reports, 14(1):24396, 2024.

[4] Z. I. Attia, P. A. Noseworthy, F. Lopez-Jimenez, S. J. Asirvatham, A. J. Deshmukh, B. J. Gersh, R. E. Carter, X. Yao, A. A. Rabinstein, B. J. Erickson, et al. An artificial intelligence-enabled ecg algorithm for the identification of patients with atrial fibrillation during sinus rhythm: a retrospective analysis of outcome prediction. The Lancet, 394(10201):861–867, 2019.

[5] N. A. Baghdadi, S. M. Farghaly Abdelaliem, A. Malki, I. Gad, A. Ewis, and E. Atlam. Advanced machine learning techniques for cardiovascular disease early detection and diagnosis. Journal of Big Data, 10(1):144, 2023.

[6] K. Battocchi, E. Dillon, M. Hei, G. Lewis, P. Oka, M. Oprescu, and V. Syrgkanis. EconML: A python package for ml-based heterogeneous treatment effects estimation. https://github.com/py-why/EconML, 2019.

[7] J. Bernett, D. B. Blumenthal, D. G. Grimm, F. Haselbeck, R. Joeres, O. V. Kalinina, and M. List. Guiding questions to avoid data leakage in biological machine learning applications. Nature Methods, 21(8):1444–1453, 2024.

[8] D. Betai, A. S. Ahmed, P. Saxena, H. Rashid, H. Patel, A. Shahzadi, A. G. Mowo-Wale, and Z. Nazir. Gender disparities in cardiovascular disease and their management: a review. Cureus, 16(5), 2024.

[9] A. Carriero, A. de Hond, B. Cappers, F. Paulovich, S. Abeln, K. G. Moons, and M. van Smeden. Explainable ai in healthcare: to explain, to predict, or to describe? Diagnostic and prognostic research, 9(1):29, 2025.

[10] Centers for Disease Control and Prevention. Fast facts: Health and economic costs of chronic conditions, June 2026. Content source: National Center for Chronic Disease Prevention and Health Promotion. Accessed: 2026-06-14.

[11] V. Chernozhukov, D. Chetverikov, M. Demirer, E. Duflo, C. Hansen, W. Newey, and J. Robins. Double/debiased machine learning for treatment and structural parameters, 2018.

[12] J. Chiha, P. Mitchell, B. Gopinath, A. J. Plant, P. Kovoor, and A. Thiagalingam. Gender differences in the severity and extent of coronary artery disease. IJC Heart & Vasculature, 8:161–166, 2015.

[13] S. Dhanka and S. Maini. A hybridization of xgboost machine learning model by optuna hyperparameter tuning suite for cardiovascular disease classification with significant effect of outliers and heterogeneous training datasets. International journal of cardiology, 420:132757, 2025.

[14] L. S. Dhingra, A. Aminorroaya, E. K. Oikonomou, A. A. Nargesi, F. P. Wilson, H. M. Krumholz, and R. Khera. Use of wearable devices in individuals with or at risk for cardiovascular disease in the us, 2019 to 2020. JAMA Network Open, 6(6):e2316634, 2023.

[15] M. Dorraki, Z. Liao, D. Abbott, P. J. Psaltis, E. Baker, N. Bidargaddi, H. R. Wardill, A. Van Den Hengel, J. Narula, and J. W. Verjans. Improving cardiovascular disease prediction with machine learning using mental health data: a prospective uk biobank study. JACC: Advances, 3(9_Part_2):101180, 2024.

[16] R. B. D’Agostino Sr, R. S. Vasan, M. J. Pencina, P. A. Wolf, M. Cobain, J. M. Massaro, and W. B. Kannel. General cardiovascular risk profile for use in primary care: the framingham heart study. Circulation, 117(6):743–753, 2008.

[17] M. Feurer and F. Hutter. Hyperparameter optimization. In Automated machine learning: Methods, systems, challenges, pages 3–33. Springer, 2019.

[18] S. D. Fihn, J. M. Gardin, J. Abrams, K. Berra, J. C. Blankenship, A. P. Dallas, P. S. Douglas, J. M. Foody, T. C. Gerber, A. L. Hinderliter, et al. 2012 accf/aha/acp/aats/pcna/scai/sts guideline for the diagnosis and management of patients with stable ischemic heart disease: a report of the american college of cardiology foundation/american heart association task force on practice guidelines, and the american college of physicians, american association for thoracic surgery, preventive cardiovascular nurses association, society for cardiovascular angiography and interventions, and society of thoracic surgeons. Journal of the American College of Cardiology, 60(24):e44–e164, 2012.

[19] L. Freijeiro-González, M. Febrero-Bande, and W. González-Manteiga. A critical review of lasso and its derivatives for variable selection under dependence among covariates. International Statistical Review, 90(1):118–145, 2022.

[20] S. Ghobrial, D. Nasiakos, N. Kinnman, W. Andrawes, H. Jackson-Smith, M. Elbouhi, A. Sarantopoulos, J. Luckmann, M. Sarkar, K. Andrawes, et al. Gap-cp: Gender, age, and presentation in chest pain. Cureus, 17(8), 2025.

[21] P. Ghosh, S. Azam, M. Jonkman, A. Karim, F. J. M. Shamrat, E. Ignatious, S. Shultana, A. R. Beeravolu, and F. De Boer. Efficient prediction of cardiovascular disease using machine learning algorithms with relief and lasso feature selection techniques. IEEE Access, 9:19304–19326, 2021.

[22] D. C. Goff, D. M. Lloyd-Jones, G. Bennett, S. Coady, R. B. D’agostino, R. Gibbons, P. Greenland, D. T. Lackland, D. Levy, C. J. O’donnell, et al. 2013 acc/aha guideline on the assessment of cardiovascular risk: a report of the american college of cardiology/american heart association task force on practice guidelines. Journal of the American College of Cardiology, 63(25 Part B):2935–2959, 2014.

[23] P. Hall and M. A. Martin. On bootstrap resampling and iteration. Biometrika, 75(4):661–671, 1988.

[24] T. Hastie, R. Tibshirani, and J. Friedman. The Elements of Statistical Learning: Data Mining, Inference, and Prediction. Springer, New York, NY, 2nd edition, 2009.

[25] A. Janosi, W. Steinbrunn, M. Pfisterer, and R. Detrano. Heart disease. UCI Machine Learning Repository, 1989. Dataset.

[26] V. V. R. Karna, V. R. Karna, V. Janamala, V. K. R. Devana, V. R. S. Ch, and A. B. Tummala. A comprehensive review on heart disease risk prediction using machine learning and deep learning algorithms. Archives of Computational Methods in Engineering, 32(3):1763–1795, 2025.

[27] R. Katarya and S. K. Meena. Machine learning techniques for heart disease prediction: a comparative study and analysis. Health and Technology, 11(1):87–97, 2021.

[28] H.-L. Kim, H.-J. Kim, M. Kim, S. M. Park, H. J. Yoon, Y. S. Byun, S. M. Park, M. S. Shin, K.-S. Hong, and M.-A. Kim. Sex differences in coronary angiographic findings in patients with stable chest pain: analysis of data from the korean women’s chest pain registry (korose). Biology of sex Differences, 13(1):2, 2022.

[29] A. Koloi, V. S. Loukas, C. Hourican, A. I. Sakellarios, R. Quax, P. P. Mishra, T. Lehtimäki, O. T. Raitakari, C. Papaloukas, J. A. Bosch, et al. Predicting early-stage coronary artery disease using machine learning and routine clinical biomarkers improved by augmented virtual data. European Heart Journal-Digital Health, 5(5):542–550, 2024.

[30] A. E. Korial, I. I. Gorial, and A. J. Humaidi. An improved ensemble-based cardiovascular disease detection system with chi-square feature selection. Computers, 13(6):126, 2024.

[31] B. C. Latha and S. C. Jeeva. Improving the accuracy of prediction of heart disease risk based on ensemble classification techniques. Informatics in Medicine Unlocked, 16:100203, 2019.

[32] T. Liu, A. J. Krentz, Z. Huo, and V. Ćurčin. Opportunities and challenges of cardiovascular disease risk prediction for primary prevention using machine learning and electronic health records: a systematic review. Reviews in Cardiovascular Medicine, 26(4):37443, 2025.

[33] X. Liu, S. C. Rivera, D. Moher, M. J. Calvert, A. K. Denniston, H. Ashrafian, A. L. Beam, A.-W. Chan, G. S. Collins, A. D. J. Deeks, et al. Reporting guidelines for clinical trial reports for interventions involving artificial intelligence: the consort-ai extension. The Lancet Digital Health, 2(10):e537–e548, 2020.

[34] A. H. Maas and Y. E. Appelman. Gender differences in coronary heart disease. Netherlands Heart Journal, 18(12):598–603, 2010.

[35] I. D. Mienye and N. Jere. Optimized ensemble learning approach with explainable ai for improved heart disease prediction. Information, 15(7):394, 2024.

[36] F. Mostafa, E. Hasan, M. Williamson, and H. Khan. Statistical machine learning approaches to liver disease prediction. Livers, 1(4):294–312, 2021.

[37] G. Nakas, A. Bechlioulis, A. Marini, K. Vakalis, M. Bougiakli, S. Giannitsi, K. Nikolaou, E. I. Antoniadou, A. Kotsia, K. Gartzonika, et al. The importance of characteristics of angina symptoms for the prediction of coronary artery disease in a cohort of stable patients in the modern era. Hellenic Journal of Cardiology, 60(4):241–246, 2019.

[38] B. Olawade, A. A. Soladoye, B. A. Omodunbi, N. Aderinto, and I. A. Adeyanju. Comparative analysis of machine learning models for coronary artery disease prediction with optimized feature selection. International Journal of Cardiology, 436:133443, 2025.

[39] J. Pepine, K. C. Ferdinand, L. J. Shaw, K. A. Light-McGroary, R. U. Shah, M. Gulati, C. Duvernoy, M. N. Walsh, C. N. Bairey Merz, and A. C. in women committee. Emergence of nonobstructive coronary artery disease: a woman’s problem and need for change in definition on angiography. Journal of the American College of Cardiology, 66(17):1918–1933, 2015.

[40] T. Pezel, S. Toupin, V. Bousson, K. Hamzi, T. Hovasse, T. Lefevre, B. Chevalier, T. Unterseeh Sanguineti, S. Champagne, et al. A machine learning model using cardiac ct and mri data predicts cardiovascular events in obstructive coronary artery disease. Radiology, 314(1):e233030, 2025.

[41] M. S. H. Rabbi, M. M. Bari, T. Debnath, A. Rahman, A. K. Das, M. P. Hossain, and G. Muhammad. Performance evaluation of optimal ensemble learning approaches with pca and lda-based feature extraction for heart disease prediction. Biomedical Signal Processing and Control, 101:107138, 2025.

[42] S. Raghunath, A. E. Ulloa Cerna, L. Jing, D. P. VanMaanen, J. Stough, D. N. Hartzel, J. B. Leader, H. L. Kirchner, M. C. Stumpe, A. Hafez, et al. Prediction of mortality from 12-lead electrocardiogram voltage data using a deep neural network. Nature medicine, 26(6):886–891, 2020.

[43] S. C. Rivera, X. Liu, A.-W. Chan, A. K. Denniston, M. J. Calvert, H. Ashrafian, A. L. Beam, G. S. Collins, A. Darzi, J. J. Deeks, et al. Guidelines for clinical trial protocols for interventions involving artificial intelligence: the spirit-ai extension. The Lancet Digital Health, 2(10):e549–e560, 2020.

[44] C. Rudin. Stop explaining black box machine learning models for high stakes decisions and use interpretable models instead. Nature machine intelligence, 1(5):206–215, 2019.

[45] J. Sen and S. Bhattacharya. An explainable hybrid framework for early detection of cardiovascular diseases using categorical boosting and bees algorithm. Scientific Reports, 2025.

[46] G. Sharma, A. E. Gaffey, A. Hameed, N. A. Kasparian, R. Mauricio, E. B. Marsh, D. Beck, J. Skowronski, D. Wolfe, G. N. Levine, and American Heart Association Women’s Health Science Committee of the Council on Clinical Cardiology and the Stroke Council. Optimizing psychological health across the perinatal period: An update on maternal cardiovascular health: A scientific statement from the american heart association. Journal of the American Heart Association, 14(5):e041369, 2025.

[47] Y. Si, A. Abdollahi, N. Ashrafi, G. Placencia, E. Pishgar, K. Alaei, and M. Pishgar. Optimized feature selection and advanced machine learning for stroke risk prediction in revascularized coronary artery disease patients. BMC Medical Informatics and Decision Making, 25(1):276, 2025.

[48] R. Singh and N. S. Mangat. Stratified sampling. In Elements of survey sampling, pages 102–144. Springer, 1996.

[49] M. A. Talukder, A. S. Talaat, and M. Kazi. Hxai-ml: a hybrid explainable artificial intelligence based machine learning model for cardiovascular heart disease detection. Results in Engineering, 25:104370, 2025.

[50] F. Tang and H. Ishwaran. Random forest missing data algorithms. Statistical Analysis and Data Mining: The ASA Data Science Journal, 10(6):363–377, 2017.

[51] J. Tanha, Y. Abdi, N. Samadi, N. Razzaghi, and M. Asadpour. Boosting methods for multi-class imbalanced data classification: an experimental review. Journal of Big data, 7(1):70, 2020.

[52] M. D. Teja and G. M. Rayalu. Optimizing heart disease diagnosis with advanced machine learning models: a comparison of predictive performance. BMC cardiovascular disorders, 25(1):212, 2025.

[53] S. Tonekaboni, S. Joshi, M. D. McCradden, and A. Goldenberg. What clinicians want: contextualizing explainable machine learning for clinical end use. In Machine learning for healthcare conference, pages 359–380. PMLR, 2019.

[54] S. Wager and S. Athey. Estimation and inference of heterogeneous treatment effects using random forests. Journal of the American Statistical Association, 113(523):1228–1242, 2018.

[55] J. Wang, Y. Xu, L. Liu, W. Wu, C. Shen, H. Huang, Z. Zhen, J. Meng, C. Li, Z. Qu, et al. Comparison of lasso and random forest models for predicting the risk of premature coronary artery disease. BMC Medical Informatics and Decision Making, 23(1):297, 2023.

[56] World Health Organization. Cardiovascular diseases (cvds). https://www.who.int/news-room/fact-sheets/detail/cardiovascular-diseases-(cvds), 2025. Fact sheet, accessed 9 March 2026.

[57] Y. Xi, H. Wang, and N. Sun. Machine learning outperforms traditional logistic regression and offers new possibilities for cardiovascular risk prediction: A study involving 143,043 chinese patients with hypertension. Frontiers in cardiovascular medicine, 9:1025705, 2022.

